# Blood flow rather than oxygen extraction accounts for a size-dependent capillary-function DSC-MRI oxygen-metabolism contrast in glioblastoma

**DOI:** 10.64898/2026.08.14.26360305

**Authors:** Marco Öchsner, Antonia Neubauer, Robert Stahl, Thomas Liebig, Robert Forbrig, Jonas Reis

## Abstract

**Background:** Dynamic susceptibility contrast MRI with capillary-function post-processing exports a relative maximum cerebral metabolic rate of oxygen, formed from blood flow and a transit-time-derived extraction term. The share each contributes to an observed contrast is unquantified in glioblastoma.

**Methods:** In a retrospective single-centre cohort with untreated glioblastoma, six perfusion maps normalised to normal-appearing white matter were sampled in automatically segmented enhancing tumour and peritumoral brain. The paired compartment contrast in the oxygen-metabolism index was partitioned into flow, extraction and residual terms and examined against tumour-core volume.

**Results:** Of 131 included patients, 122 were suitable for analysis. Flow-linked maps were about twice as high in enhancing tumour, with transit and extraction maps only modestly increased (all q < 0.05). Flow accounted for 92.6% (95% CI 85.9–98.8) of the contrast and extraction for 6.6% (0.7–12.9). Across volume tertiles the flow share rose from 67.8% to 104.0%, a gradient arising peritumorally: every map changed with volume there, none in enhancing tumour.

**Conclusion:** The compartment contrast in the oxygen-metabolism index is largely accounted for by blood flow and varies with lesion size, that dependence originating peritumoral tissue. It should be interpreted within the complete perfusion panel, not as independent metabolic evidence.

## Introduction

In isocitrate dehydrogenase (*IDH*)-wildtype glioblastoma, relative cerebral blood volume (rCBV) from dynamic susceptibility contrast (DSC) MRI is the most standardised perfusion biomarker and is routinely interpreted alongside the contrast-enhancing (CE) component and the surrounding non-enhancing T2/fluid-attenuated inversion recovery (FLAIR) abnormality, the peritumoral brain zone (PBZ) [1, 2, 3]. Capillary-function modelling of the same acquisition additionally exports mean transit time (MTT), capillary transit time heterogeneity (CTH) and a maximum oxygen extraction fraction (OEF^max^) derived from the fitted transit-time distribution. It also exports an index of the relative maximal cerebral metabolic rate of oxygen (rCMRO₂^max^), formed as the product of OEF^max^ and relative cerebral blood flow (rCBF) [4, 5].

These maps are increasingly reported as indices of tumour metabolism: DSC-derived CTH and OEF^max^ have been used to separate pseudoprogression from true progression after treatment [6], lower-tail rCMRO₂ and OEF^max^ to predict *IDH* mutation [7] and overall survival in *IDH*-wildtype glioblastoma [8].

The extent to which observed rCMRO₂^max^ differences reflect changes in flow or in oxygen extraction, and whether that balance is stable across lesions of different size, has not to our knowledge been established for this model family. The same model family has been applied to tumour oxygenation in recurrent bevacizumab-treated glioblastoma without quantifying the component shares [9], and region-of-interest sampling established that an enhancing-versus-peritumoral difference exists in untreated disease [10] without the whole-compartment features required to determine its composition.

The model fixes how flow and extraction combine within the index, but not how much of an observed contrast each carries, and that division has not been quantified in tumour. We therefore partitioned the enhancing-versus-peritumoral rCMRO₂^max^ contrast into its flow, extraction and residual terms, characterised the accompanying six-map perfusion phenotype, and assessed whether the contrast and its component balance vary with tumour-core volume. Compartments were sampled by automated whole-compartment segmentation, an approach established as an effective alternative to focal region-of-interest sampling [11] and, in untreated glioblastoma, more reproducible than hot-spot sampling [12].

## Methods

This retrospective study was approved by the institutional review board of LMU Munich (approval no. 24-0692), which waived the requirement for written informed consent, and was conducted in accordance with the Declaration of Helsinki.

### Cohort

We performed a single-centre retrospective cross-sectional study of adults with newly diagnosed, untreated *IDH*-wildtype glioblastoma (2021 WHO classification) imaged in routine care between January 2020 and July 2025. Consecutive pathology-confirmed cases were identified by systematic database query, linked to the imaging archive, and screened against the imaging criteria. No sample-size calculation was performed, and the study sample size was determined by the number of eligible cases within the accrual window. Patient selection is detailed in Supplementary Figure S1. The study cohort comprised 131 patients, of whom 122 had both compartments available for the paired analysis; under pairwise deletion the analysed number differs by endpoint (122 perfusion, 120 ADC; Supplementary Table S1). Sixty-eight of the 131 patients were also included in a prior region-of-interest report (n = 72) [10] and 74 in a second region-of-interest study relating peritumoral oxygen extraction to Ki-67 (n = 80) [13]. Neither prior report used whole-compartment segmentation or partitioned the contrast.

### Imaging and maps

MRI was performed on 1.5-T (Magnetom Sola Fit, Siemens Healthineers) and 3-T scanners (Signa HD and Signa Premier, GE Healthcare; Magnetom Vida, Siemens Healthineers) using the institutional protocol, with a preload-based DSC acquisition using gadobutrol (Gadovist, Bayer) (Supplementary Methods and Supplementary Table S1). DSC data were processed with vendor-agnostic capillary-function software (Neurosuite v16.2, release 2023-02-01-03; Cercare Medical) providing rCBV, rCBF, MTT, CTH, OEF^max^ and rCMRO₂^max^. A single global arterial input function was selected automatically and applied voxelwise. Each voxel concentration–time curve was fitted with a physiologically constrained gamma residue using Bayesian expectation–maximisation rather than singular-value-decomposition deconvolution [14], updating CBF, MTT, macrovascular delay and noise variance; a unidirectional leakage term was co-estimated within the voxelwise model, yielding leakage-corrected maps [5, 15]. CBV is analytically equal to CBF × MTT in the continuous model, but the exported CBV map is estimated separately by numerical integration of the fitted leakage-corrected tissue concentration curve divided by the arterial-input-function area, so exported CBV does not equal the product identically (Supplementary Methods).

### Map hierarchy

The exported maps form a structured perfusion system rather than independent measurements. rCBF, MTT and CTH are the fitted transport coordinates: flow, mean transit time and transit-time dispersion, jointly estimated from the residue model. rCBV, OEF^max^ and rCMRO₂^max^ are model-linked or derived outputs: rCBV is model-linked to rCBF × MTT, as described above; OEF^max^ is a model-derived extraction-capacity index obtained from the fitted transit-time distribution, of which MTT and CTH are the mean and standard deviation, under fixed software assumptions; and rCMRO₂^max^ is proportional to OEF^max^ × rCBF. ADC, an independent diffusion measure, is reported as supportive. The model and its exported quantities are described elsewhere [4, 5, 16], with glioma applications [7, 8] and a comparison with oxygen-15 PET in healthy brain [17]; Supplementary Methods give the patient-specific normalisation constant that cancels in the within-patient log difference and the closure assessment for the rCMRO₂^max^ relation.

### Segmentation and compartments

Automated segmentation used a T2-FLAIR-dropout nnU-Net model [18], yielding CE tumour, necrosis, and oedema/T2-FLAIR abnormality; tumour core = CE + necrosis, PBZ = T2/FLAIR-abnormal tissue outside core. Necrosis was characterised descriptively only, and because the segmentation does not separate infiltrative tumour from oedema the comparator was PBZ abnormality (Supplementary Methods). All continuous parametric maps were rigidly registered and resampled once into each patient’s post-contrast T1-weighted space using trilinear interpolation; segmentation and parcellation labels were resampled by nearest neighbour. All primary analyses used normal-appearing white matter (NAWM)-normalised maps. The reference was an automatically generated contralateral cerebral white-matter mask; for tumours crossing the midline it was the tumour-free anterior or posterior white-matter third. All voxels within 10 mm of the whole tumour segmentation were excluded from the reference (Supplementary Methods, Registration and NAWM normalisation). Unless stated otherwise, all reported map values are NAWM-normalised. The prefix r is retained only for the relative perfusion outputs rCBV, rCBF and rCMRO₂^max^. Cross-patient feature summaries were harmonised for scanner using location-only ComBat (vendor × field strength), to which the paired contrasts and the decomposition are invariant (Supplementary Methods, Harmonisation) [19, 20]. A layered quality-control framework preceded analysis (Supplementary Methods, Quality control): WARN cases were retained in the primary analyses and excluded in a PASS-only sensitivity analysis.

### Features

A histogram-based feature set was used. The NAWM-normalised maps were positive and right-skewed, so the natural logarithm of the normalised value was the primary analytic scale, and effect sizes are reported as back-transformed fold-changes. Non-positive values occurred only within necrosis, which was excluded from inferential analyses and retained only for descriptive quality-control assessment. For each compartment and map we extracted the median, the interquartile range, upper-tail summaries and the geometric-mean feature required by the decomposition. The geometric mean is the only summary that is linear over the voxel logarithms, so it is the summary for which the model identity is additive within a patient, while the median and upper-tail features are reported for the compartment. comparisons but do not satisfy that identity. Features were computed on finite, strictly positive voxels within each map, and upper-tail features were computed only for compartments exceeding a minimum analysable volume, further information given in in Supplementary Methods, Feature extraction.

### Statistics

Distributional assumptions were assessed by Shapiro–Wilk tests. On the ratio scale six of seven paired compartment contrasts departed from normality; while on the natural-log scale two did. Non-parametric and distribution-free methods were therefore used throughout, and regression estimates use heteroscedasticity-consistent (HC3) standard errors, which do not assume normally distributed residuals. Compartment contrasts were tested for the six perfusion maps together — rCBV, rCBF, MTT, CTH, OEF^max^ and rCMRO₂^max^ — as a median-feature family (m = 6) and a distributional family (m = 22): rCBV, rCBF, MTT, CTH, OEF^max^ and rCMRO₂^max^. This grouping is inferential, and does not imply that the six maps are statistically or physiologically independent (two-sided Wilcoxon signed-rank tests on paired log-differences; Hodges–Lehmann pseudomedians of the paired log-differences, back-transformed to fold-changes with bias-corrected accelerated (BCa) bootstrap 95% CIs). ADC was reported as a supportive non-perfusion comparison and was not entered into either correction. Benjamini–Hochberg correction was applied within each family (28 corrected endpoints; Supplementary Table S2). The endpoint registry and principal analysis procedures were documented in an internal analysis specification (8 July 2026) before the final analysis run (22 July 2026). Analyses not contained in that specification are identified as exploratory. Table 2 reports the full perfusion panel. rCBF and MTT are interpreted with the caveat that they are more sensitive to arterial-input-function selection than rCBV.

### Decomposition

OEF^max^ is determined by the transit-time distribution alone, carries no patient-specific tissue-oxygen-tension term, and is an upper bound on extraction. Within the model rCMRO₂^max^ is proportional to OEF^max^ × rCBF. After separate normalisation of each map to a common NAWM reference the relation holds up to a patient-specific constant, because the NAWM reference statistics need not themselves satisfy the product identity; that constant is common to both compartments and cancels in the within-patient CE-versus-PBZ log difference. For the rCMRO₂^max^ module, the contrast was partitioned using per-compartment geometric-mean features, the only summary on which the additive identity holds: Δlog rCMRO₂^max^ = Δlog rCBF + Δlog OEF^max^ + ε. The primary decomposition is the mean-of-logs partition, which holds exactly because the identity is additive within patients; each component’s share of the mean contrast is reported with BCa bootstrap intervals over patients (Supplementary Methods). To determine which compartment generates the lesion-size dependence of the contrast, each compartment-versus-NAWM contrast was regressed on log₂ tumour-core volume by ordinary least squares with heteroscedasticity-consistent (HC3) intervals, on the same geometric-mean features; the absolute NAWM reference was regressed on the same predictor as a stability check. Separately, Δlog rCMRO₂^max^ was regressed on Δlog rCBV (robust regression, with ordinary least squares alongside). Because both are model-linked to flow, this quantifies their association rather than testing physiological independence.

### Software

Segmentation, co-registration, NAWM mask generation, feature extraction and statistical analyses were performed in Python 3.13.5 (NumPy 2.4.4, pandas 3.0.2, SciPy 1.17.1, scikit-learn 1.6.1, nibabel 5.3.2, statsmodels 0.14.6) using ANTs 2.6.5, SynthSeg 2.0 and nnU-Net v2 (2.6.2), and figures generated with matplotlib 3.6.3 and SciencePlots 2.2.0.

## Results

### Cohort

The final study cohort comprised 131 patients, of whom 122 were included for the paired comparisons after the 0.5 mL inclusion floor. Cohort characteristics, acquisition context, and lesion morphology are summarised in Table 1; the derivation is shown in Supplementary Figure S1; with representative cases are shown in Figure 1. NAWM normalisation passed quality control in all included cases.

**Figure 1.**
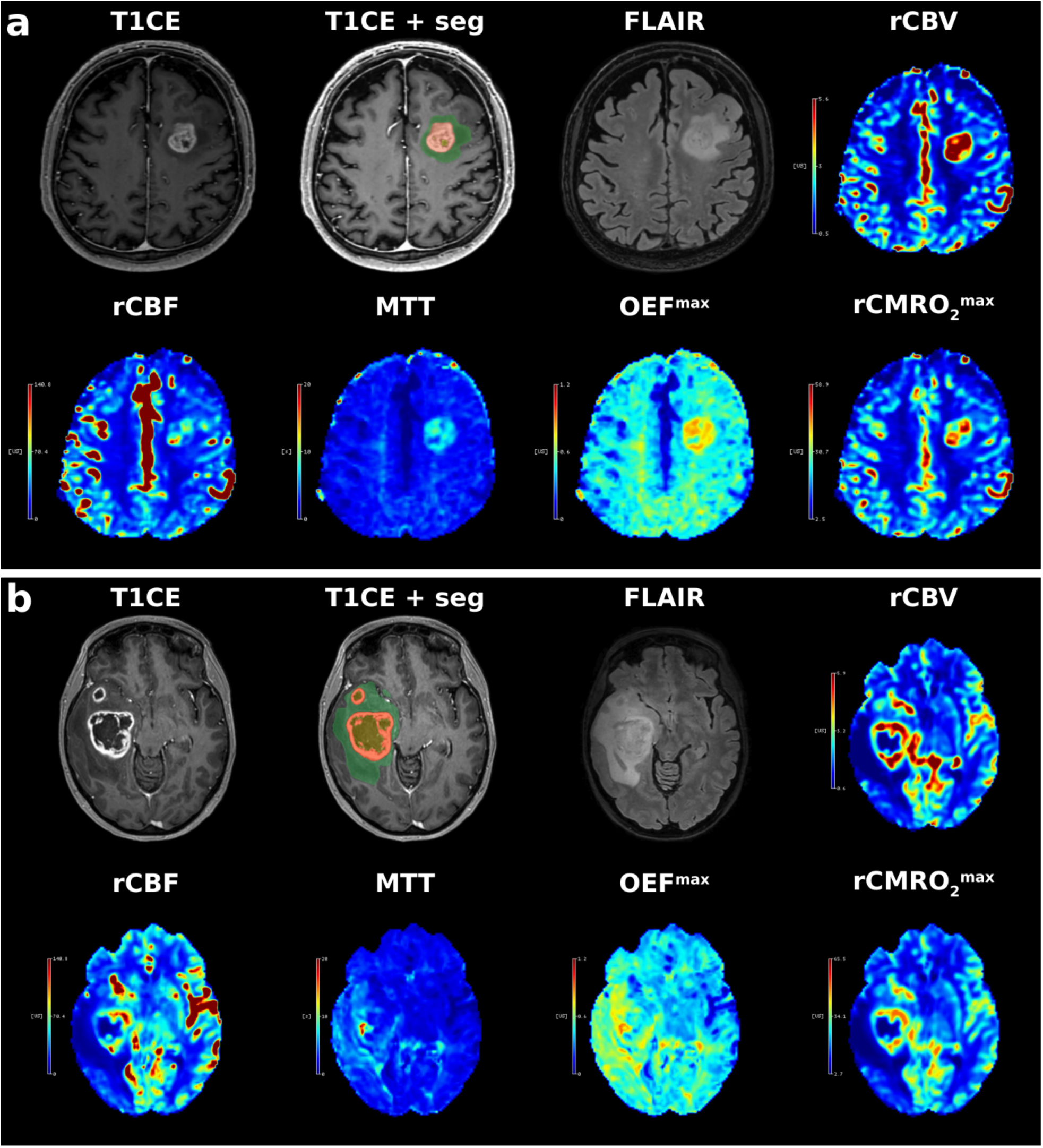
Compartment segmentation and capillary-function perfusion maps in two patients. Both cases passed all quality-control domains, contained a single lesion and did not cross the midline. (a) A 9.8 mL tumour core with minimal necrosis (0.4 mL). (b) A 39.5 mL core with substantial necrosis (12.3 mL) and a larger peritumoral zone. For each patient one co-registered axial slice at the level of maximal contrast-enhancing area is shown: post-contrast T1-weighted without and with the segmentation overlaid, then T2/FLAIR, rCBV, rCBF, MTT, OEF^max^ and rCMRO₂^max^. The segmentation panel is the only panel carrying an overlay: contrast-enhancing tumour in orange, necrosis in olive and peritumoral brain zone in green. Each map has its own colour bar, identical for both patients; scales are not shared across maps. Case (a) shows prolonged enhancing-relative transit with correspondingly elevated OEF^max^, a pattern not typical of the cohort (median ratio ×1.05, Table 2). Case (b) shows the more usual configuration, in which the enhancing rim is conspicuous on rCBV and rCBF while MTT, OEF^max^ and rCMRO₂^max^ differ less between compartments. FLAIR, fluid-attenuated inversion recovery; MTT, mean transit time; OEF^max^, maximum oxygen extraction fraction; rCBF, relative cerebral blood flow; rCBV, relative cerebral blood volume; rCMRO₂^max^, relative maximum cerebral metabolic rate of oxygen.

**Table 1.** Cohort characteristics, acquisition context, and lesion morphology (n = 131). Compartment volumes are summarised over patients in whom the label was present; patients without an enhancing or necrotic label contribute no value rather than zero. Multiple lesions denotes more than one spatially disconnected tumour-core component on connected-component analysis, confirmed by visual review; maximal diameter refers to the largest core component in such cases.

| Characteristic | Value |
| --- | --- |
| Age, years | 67.9 [58.7, 77.4] |
| Sex, female / male | 51 / 80 |
| Vendor × field: GE 3 T / Siemens 3 T / Siemens 1.5 T | 71 / 35 / 25 |
| Tumour core volume, mL | 15.4 [5.7, 37.1] |
| Enhancing (CE) volume, mL | 13.3 [5.7, 27.2] |
| Peritumoral (PBZ) volume, mL | 36.6 [15.2, 72.3] |
| Necrosis volume, mL | 3.2 [0.6, 10.2] |
| Tumour core maximal diameter, mm | 30.8 [22.2, 41.4] |
| Multiple lesions, n (%) | 13 (9.9) |
| Whole tumour crosses midline, n (%) | 48 (36.6) |

### Compartment contrasts

The six perfusion maps separated into two magnitude groups. CE tumour showed markedly higher rCBV (×2.38 [95% CI 2.14–2.62]), rCBF (×2.06 [1.79–2.36]) and rCMRO₂^max^ (×2.14 [1.93–2.38]) than PBZ, and smaller but directionally coherent increases in MTT (×1.13 [1.07–1.20]), CTH (×1.15 [1.10–1.21]) and OEF^max^ (×1.05 [1.01–1.08]); all six met q < 0.05 (Table 2 and Figure 2). ADC was lower in CE as a supportive non-perfusion comparison (×0.93 [0.90–0.96]; Supplementary Table S5). The distributional endpoints followed the same directions, with smaller upper-tail than median effects (rCBV P90 ×1.76 [1.63, 1.89]) and lower relative within-compartment dispersion in CE for the three flow-linked maps (Supplementary Table S4). Absolute OEF^max^ was well below its upper bound in every compartment (medians 0.46 CE, 0.44 PBZ, 0.43 NAWM; Supplementary Table S5). The geometric-mean features used for the decomposition give slightly smaller contrasts (rCMRO₂^max^ ×1.99, Table 3). All six effects were stable in the segmentation PASS-only subset, under a stricter volume floor and without harmonisation (Supplementary Table S4), and across acquisition strata and structural input configurations (Supplementary Table S4). Point estimates for OEF^max^ were larger and those for CTH smaller at 1.5 T, although the small subgroup (n = 24) and overlapping intervals preclude inference about effect modification.

**Figure 2.**
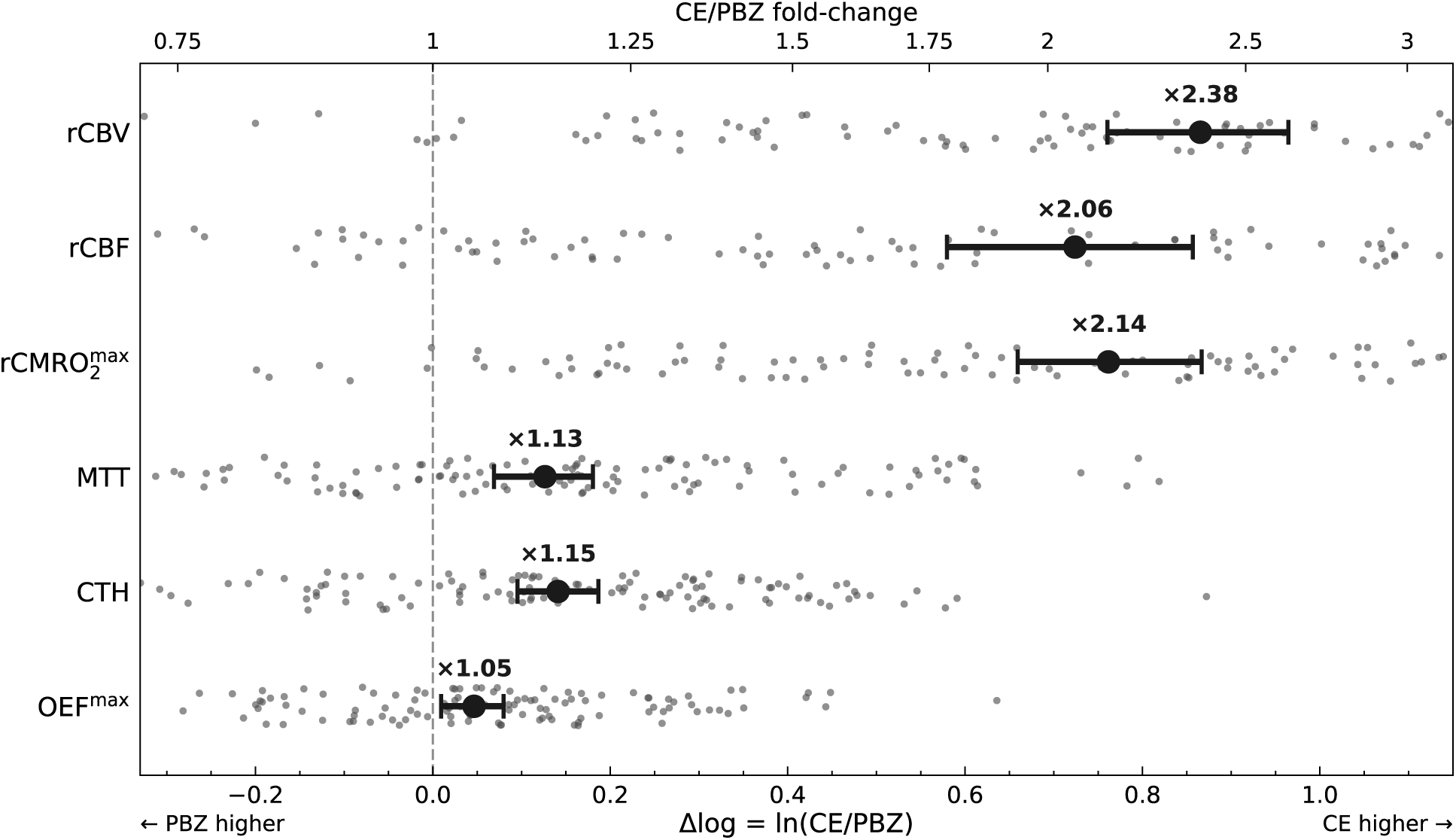
Paired CE-versus-PBZ compartment effects for the perfusion panel. For each patient and map the within-patient log-difference Δlog = ln(CE/PBZ) in the NAWM-normalised compartment median is plotted (small points; positive values indicate higher CE); the bold marker and horizontal bar give the Hodges–Lehmann paired estimate with its bias-corrected accelerated (BCa) bootstrap 95% confidence interval (10,000 resamples), and the back-transformed CE/PBZ fold-change is labelled (upper axis). The six perfusion maps of the median-feature family are shown, ordered as two magnitude clusters. All six differed between compartments after Benjamini–Hochberg correction within the family (two-sided Wilcoxon signed-rank tests; q < 0.05 throughout, largest q = 0.018 for OEF^max^; Table 2). Effects are computed on median features; the geometric-mean features used in the flow–extraction decomposition (Figure 3) yield slightly smaller contrasts. rCBF and MTT depend on arterial-input-function selection; compartment-effect directions were consistent across acquisition strata (Supplementary Table S4). n = 122. ADC, apparent diffusion coefficient; CE, contrast-enhancing tumour; CTH, capillary transit time heterogeneity; MTT, mean transit time; OEF^max^, maximum oxygen extraction fraction; NAWM, normal-appearing white matter; PBZ, peritumoral brain zone; rCBF, relative cerebral blood flow; rCBV, relative cerebral blood volume; rCMRO₂^max^, relative maximum cerebral metabolic rate of oxygen.

**Table 2.**
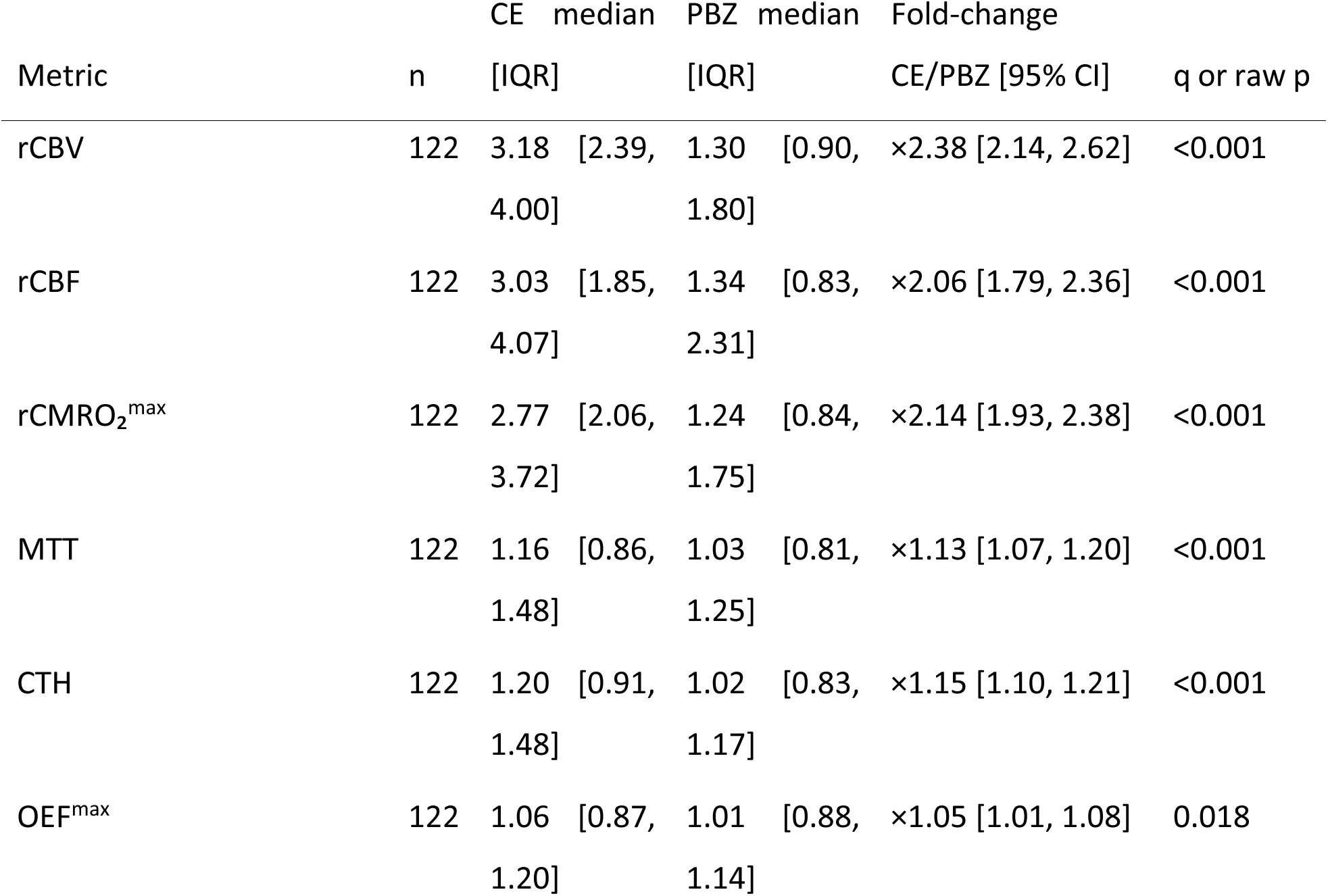
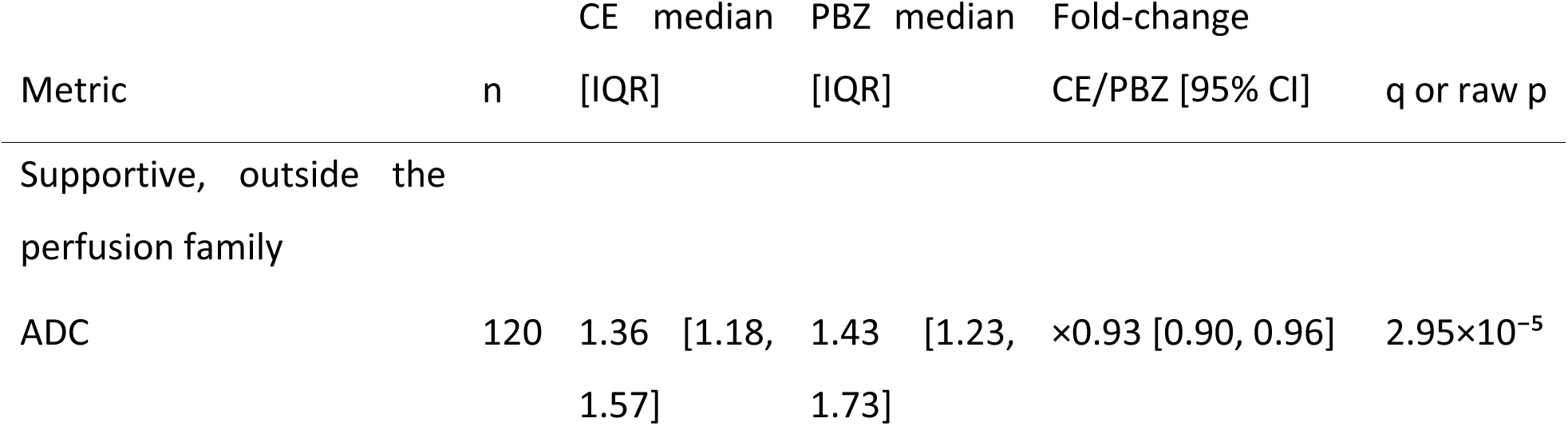
Six-map CE-versus-PBZ perfusion phenotype and supportive ADC comparison. Effects are Hodges–Lehmann estimates of the paired CE-minus-PBZ log-difference, back-transformed to a fold-change, with BCa 95% CIs (10,000 bootstrap resamples). Rows are ordered by effect magnitude, the three flow-linked maps first. The six perfusion maps form the median-feature family, with q from Benjamini–Hochberg correction within that family; ADC is reported as a supportive non-perfusion comparison; it was not entered into the multiplicity correction, and the value shown in the final column is a raw p value. rCBF and MTT are more sensitive to arterial-input-function selection than rCBV, but their compartment effects were directionally consistent across acquisition strata (Supplementary Table S4). n differs because two patients lacked diffusion data.

**Table 3.** Flow–extraction decomposition of the CE-versus-PBZ rCMRO₂^max^ contrast, pooled and by tumour-core volume tertile. Terms are arithmetic means of the patient-level log differences on per-compartment geometric-mean features, the summary for which the identity Δlog rCMRO₂^max^ = Δlog rCBF + Δlog OEF^max^ + ε₂ holds within each patient. Terms are given before shares because shares become unstable when the total contrast is smaller. Shares are the component mean divided by the total mean and may exceed 100% or be negative. Intervals are given for every term and every share, and are bias-corrected accelerated bootstrap intervals over patients (10,000 resamples, seed 20260709). The closure residual is a processing diagnostic, not a physiological component; its magnitude on the vendor-export grid is given in Supplementary Table S6. Panel C repeats the pooled decomposition in the segmentation PASS-only subset. The extraction-term intervals in the middle and largest tertiles, and in the PASS-only subset, include zero. Tertile analyses were specified as exploratory for the original compartment endpoints; their extension to the component terms is exploratory. Shares are computed on per-compartment geometric-mean features and are not reproducible from the median-feature fold-changes in Table 4, which summarise the same contrasts on a different scale. The relationship between the rCMRO₂^max^ and rCBV contrasts is shown in Supplementary Figure S2.

| Quantity | Estimate [95% CI] |
| --- | --- |
| *Panel A — pooled decomposition (n = 122)* |  |
| Total, $\Delta\log \text{rCMRO}_2^{\text{max}}$ | +0.689 [+0.590, +0.785] |
| Flow term, $\Delta\log \text{rCBF}$ | +0.638 [+0.516, +0.761] |
| Extraction term, $\Delta\log \text{OEF}^{\text{max}}$ | +0.046 [+0.006, +0.082] |
| Closure residual, $\epsilon_2$ | +0.005 [−0.003, +0.014] |
| Flow share, % | 92.6 [85.9, 98.8] |
| Extraction share, % | 6.6 [0.7, 12.9] |
| Closure residual share, % | 0.8 [−0.4, 2.2] |
| *Panel B — by tumour-core volume tertile* |  |
| *Tertile 1, smallest (n = 41; median 4.3 mL)* |  |
| Total, $\Delta\log \text{rCMRO}_2^{\text{max}}$ | +0.405 [+0.226, +0.551] |
| Flow term, $\Delta\log \text{rCBF}$ | +0.274 [+0.089, +0.455] |
| Extraction term, $\Delta\log \text{OEF}^{\text{max}}$ | +0.109 [+0.046, +0.165] |
| Flow share, % | 67.8 [38.4, 85.5] |
| Extraction share, % | 26.8 [9.9, 54.2] |
| *Tertile 2, middle (n = 40; median 17.2 mL)* |  |
| Total, $\Delta\log \text{rCMRO}_2^{\text{max}}$ | +0.746 [+0.586, +0.917] |
| Flow term, $\Delta\log \text{rCBF}$ | +0.687 [+0.480, +0.916] |
| Extraction term, $\Delta\log \text{OEF}^{\text{max}}$ | +0.057 [−0.023, +0.124] |
| Flow share, % | 92.0 [80.0, 102.6] |
| Extraction share, % | 7.6 [−2.7, 19.3] |
| *Tertile 3, largest (n = 41; median 49.5 mL)* |  |
| Total, $\Delta\log \text{rCMRO}_2^{\max}$ | +0.916 [+0.782, +1.061] |
| Flow term, $\Delta\log \text{rCBF}$ | +0.953 [+0.784, +1.127] |
| Extraction term, $\Delta\log \text{OEF}^{\max}$ | -0.028 [-0.090, +0.026] |
| Flow share, % | 104.0 [97.3, 110.6] |
| Extraction share, % | -3.1 [-9.4, 3.0] |
| *Panel C — segmentation PASS-only subset (n = 106)* |  |
| Total, $\Delta\log \text{rCMRO}_2^{\max}$ | +0.667 [+0.562, +0.767] |
| Flow term, $\Delta\log \text{rCBF}$ | +0.620 [+0.493, +0.748] |
| Extraction term, $\Delta\log \text{OEF}^{\max}$ | +0.043 [-0.001, +0.083] |
| Flow share, % | 93.1 [85.5, 99.9] |
| Extraction share, % | 6.5 [-0.0, 13.7] |

### Flow–extraction decomposition

On the geometric-mean feature used for the additive decomposition, flow constituted 92.6% (95% CI 85.9–98.8) of the mean log-scale contrast; the corresponding shares were 6.6% (0.7–12.9) for the model-based extraction ceiling and 0.8% (−0.4 to 2.2) for the closure residual (Table 3 and Figure 3). The compartment contrast in rCMRO₂^max^ closely tracked that in rCBV, which is model-linked to flow through the same fitted transport state (Supplementary Figure S2). The identity did not close exactly, but in 120 patients analysed with identical masks and inclusion rules on both grids the median absolute residual fell by 94.6% on the vendor-export grid, and was too small to alter the component shares (Supplementary Table S6).

**Figure 3.**
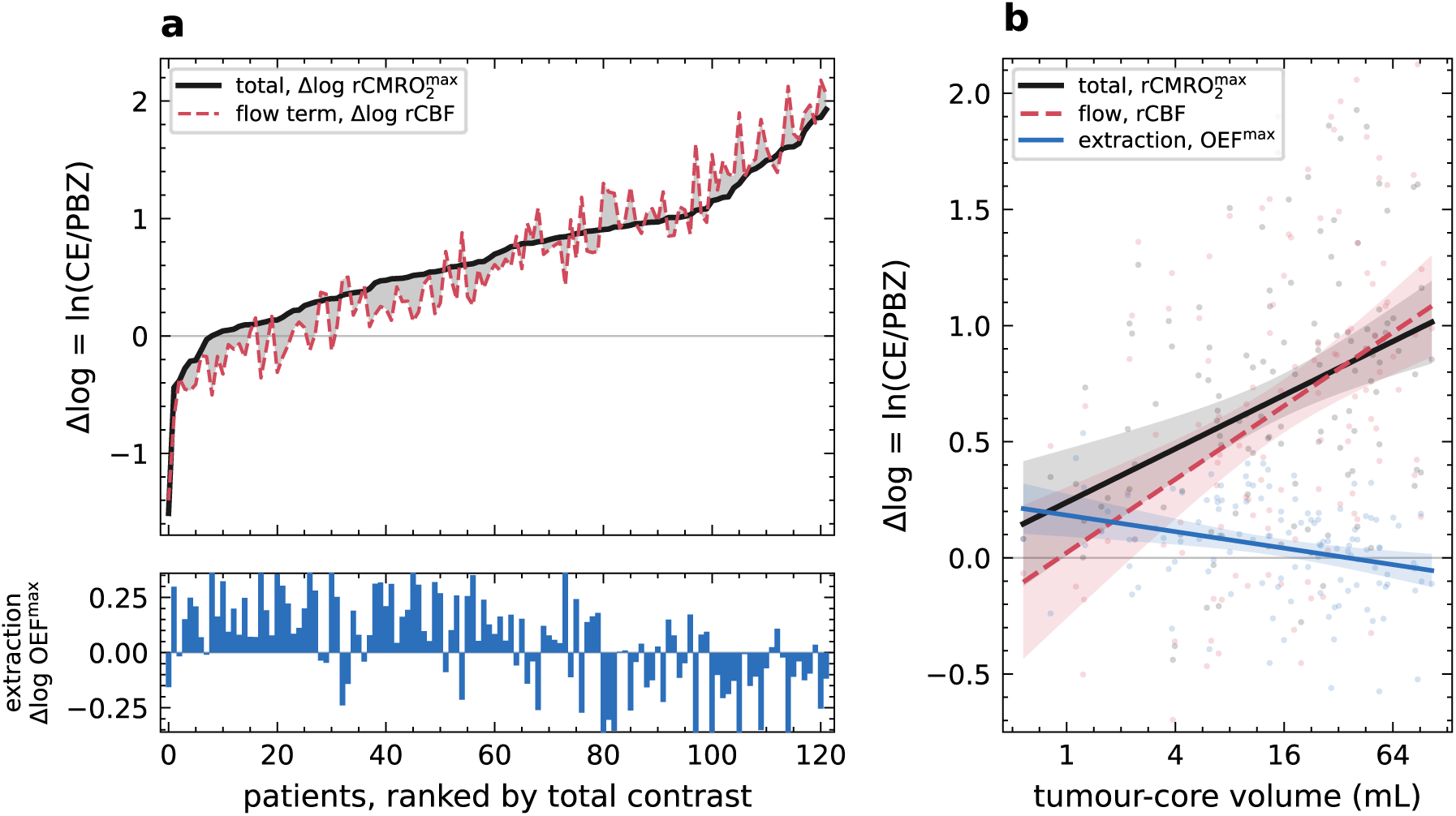
Flow–extraction decomposition of the CE-versus-PBZ rCMRO₂^max^ contrast and its dependence on lesion size (per-compartment geometric-mean features; n = 122). (a) Patients ranked by the total contrast Δlog rCMRO₂^max^ (solid line), with the flow term Δlog rCBF dashed in red; the shaded band between them is the extraction term together with the small closure residual. The strip beneath shows the extraction term alone (Δlog OEF^max^, blue), which is small in both directions: 46 of 122 patients had a negative extraction term, a feature the cohort mean share of 6.6% does not convey. (b) Flow, extraction and total contrasts against tumour-core volume, with ordinary-least-squares fits and 95% mean-prediction bands; total in black, flow in red, extraction in blue as in (a). The flow term rises by +0.158 log units per doubling of volume (95% CI +0.102 to +0.215) while the extraction term falls by −0.035 (−0.057 to −0.014); the flow line crossing the total line corresponds to the flow share exceeding 100%, which it does in the largest volume tertile (104.0%; Table 3). Bulk compartment masks cannot distinguish a biological difference, smaller lesions being less neovascularised, from a geometric one, since proportionally larger boundary regions compress the flow contrast in smaller lesions; the gradient should not be read as evidence for either. Component terms and shares with bootstrap confidence intervals are given in Table 3; the decomposition is descriptive and no significance test is attached to the shares. The volume slopes in (b) are from ordinary least squares with heteroscedasticity-consistent (HC3) intervals and both differ from zero (p = 6.1×10⁻⁶ for flow, p = 0.0015 for extraction). Δlog = ln(CE/PBZ); CE, contrast-enhancing tumour; OEF^max^, maximum oxygen extraction fraction; PBZ, peritumoral brain zone; rCBF, relative cerebral blood flow; rCMRO₂^max^, relative maximum cerebral metabolic rate of oxygen.

### Lesion size

Per doubling of tumour-core volume no perfusion map changed significantly within CE tumour (rCBF ×1.002 [95% CI 0.941, 1.066]; rCBV ×1.029 [0.990, 1.069]; rCMRO₂^max^ ×1.021 [0.978, 1.065]), whereas every perfusion map changed in the PBZ, with lower flow (rCBF ×0.855 [0.804, 0.910]; rCBV ×0.918 [0.881, 0.958]) and longer, more heterogeneous transit (MTT ×1.087 [1.047, 1.129]; CTH ×1.060 [1.034, 1.087]) (Table 4). In absolute units no NAWM map changed with tumour-core volume (rCBF ×1.017 [0.995, 1.039]; Table 4), so the compartment estimates are not a normalisation artefact. ADC rose in both compartments (CE ×1.047 [1.027, 1.068]; PBZ ×1.051 [1.033, 1.068]) with an unchanged contrast (×0.997 [0.980, 1.014]), so the divergence between compartments is perfusion-specific and peritumoral. The CE-versus-PBZ contrast is the difference of these two slopes and therefore widened for the flow-linked maps and narrowed for the transit and extraction maps as lesions grew. The decomposition moved with it: across tumour-core volume tertiles the flow term carried 67.8%, 92.0% and 104.0% of the contrast, and the extraction term fell from +0.109 to −0.028, a value indistinguishable from zero (−0.035 per doubling, 95% CI −0.057 to −0.014). Shares were unchanged in the segmentation PASS-only subset (n = 106, flow 93.1%, Table 3). Exploratory continuous estimates and robustness analyses are given in Supplementary Table S3.

**Table 4.** Change in each compartment-versus-NAWM contrast and in the CE-versus-PBZ contrast per doubling of tumour-core volume. Multiplicative change per doubling, from ordinary least-squares regression of each paired log contrast on log₂ tumour-core volume with heteroscedasticity-consistent (HC3) 95% confidence intervals, on per-compartment geometric-mean features. The CE-versus-PBZ column is the CE column divided by the PBZ column and is the only column independent of the reference. Regressed on tumour-core volume in absolute units the NAWM reference itself did not change (rCBF ×1.017 [0.995, 1.039]; rCBV ×1.003 [0.974, 1.034]; MTT ×0.985 [0.955, 1.017]; CTH ×0.996 [0.967, 1.026]; OEF^max^ ×0.987 [0.968, 1.005]; rCMRO₂^max^ ×1.002 [0.977, 1.028]). n = 122; ADC n = 120. Asterisks mark estimates whose confidence interval excludes 1. †ADC is a supportive non-perfusion comparison and was not entered into the multiplicity correction. This is an exploratory analysis. Tertile-stratified effects are given in Supplementary Table S3.

| Map | CE – NAWM | PBZ – NAWM | CE – PBZ |
| --- | --- | --- | --- |
| rCBV | $\times 1.029$ [0.990, 1.069] | $\times 0.918$ [0.881, 0.958]* | $\times 1.120$ [1.071, 1.172]* |
| rCBF | ×1.002 [0.941, 1.066] | ×0.855 [0.804, 0.910]* | ×1.171 [1.106, 1.240]* |
| rCMRO<sub>2</sub><sup>max</sup> | ×1.021 [0.978, 1.065] | ×0.909 [0.870, 0.951]* | ×1.122 [1.074, 1.174]* |
| MTT | ×1.035 [0.983, 1.090] | ×1.087 [1.047, 1.129]* | ×0.952 [0.922, 0.983]* |
| CTH | ×1.023 [0.983, 1.065] | ×1.060 [1.034, 1.087]* | ×0.965 [0.941, 0.990]* |
| OEF<sup>max</sup> | ×1.023 [0.987, 1.060] | ×1.060 [1.028, 1.092]* | ×0.965 [0.944, 0.987]* |
| ADC† | ×1.047 [1.027, 1.068]* | ×1.051 [1.033, 1.068]* | ×0.997 [0.980, 1.014] |

## Discussion

Partitioning the enhancing-versus-peritumoral rCMRO₂^max^ contrast showed that flow accounted for the great majority of the mean log difference and the model-based extraction ceiling for a small remainder. The flow-linked maps separated the compartments strongly, while transit time, its heterogeneity and the derived extraction ceiling differed only modestly. That division was not constant: the flow share rose with tumour-core volume until it exceeded the total contrast, and the extraction term fell to a value indistinguishable from zero. Resolving each contrast into its compartment-versus-NAWM components showed that this gradient arises in the peritumoral zone, where every perfusion map changed with volume, and not within enhancing tumour, where none did. We are not aware of a study quantifying the proportion of an observed contrast assigned to each component.

rCMRO₂^max^ is a derived flow–extraction composite rather than an independent measurement. The model fixes how the components combine but places no constraint on their relative size, so the share each carries of an observed contrast is an empirical quantity; the present estimates are specific to this cohort, implementation and comparison. The compartment contrast is not uniformly flow-dominated: in the smallest lesions the extraction term contributed materially, so where flow and extraction diverge the composite carries information the flow map alone does not. Whether voxels with concordant or discordant flow and extraction differ in their association with tumour biology or outcome was not examined here. The small extraction term is not a consequence of the index approaching its upper bound: absolute OEF^max^ lay well below that bound in every compartment, and the observed compartment shift used only a small fraction of the log distance remaining to it. Upper-ceiling saturation therefore does not explain the result. A second consideration is that OEF^max^ is a saturating function of the fitted transit-time distribution and therefore may compresses its dynamic range relative to rCBF. The decomposition quantifies the contribution carried by exported rCBF and OEF^max^ maps, accordingly,we cannot claim that tissue extraction is physiologically invariant.

Neither the exported extraction map nor the metabolic-rate index has, to our knowledge, been validated against an independent reference standard in tumour; even a PET-calibrated extraction model correlates only moderately with oxygen-15 PET extraction in healthy brain, over a compressed dynamic range [17]. Two single-institution studies using the same model nonetheless reported that this index carries molecular and prognostic information: the tumour 10th percentile was retained in a model selecting for *IDH* mutation [7] and, in *IDH*-wildtype glioblastoma, for overall survival, where blood volume was not [8]. Those analyses use whole-tumour masks summarised by distributional features. The present decomposition does not explain those associations: it applies to geometric-mean features, and a lower-tail summary of a product is not the product of the lower-tail summaries, so the specific features used in those studies were not decomposed here.

The size dependence constrains how the pooled estimate should be read, and its origin is one-sided. In the largest lesions the exported index behaves almost entirely as a scaled flow map; while in the smallest, the extraction term contributes about a quarter of the contrast. That change is generated by the peritumoral compartment: with increasing tumour-core volume the PBZ showed lower flow and blood volume and longer, more heterogeneous transit, while no perfusion map changed within enhancing tumour and the NAWM reference was stable in absolute units. The diffusion contrast was unchanged although diffusion rose in both compartments, which does not support uniform dilution of all compartment contrasts in smaller lesions. Two readings remain open. Peritumoral perfusion may genuinely fall as tumours enlarge, or a fixed-width peritumoral mask around a larger core may sample progressively more distant, more normal-appearing tissue; bulk masks cannot separate them, and no causal inference about angiogenesis is warranted. Fixed-distance sampling would distinguish the two, and predicts that the dependence should attenuate once distance from the enhancing edge is held constant.

This study has several limitations. First, the design was retrospective and single-centre, with residual protocol and scanner effects despite locked post-processing. Second, OEF^max^, CTH and rCMRO₂^max^ are model-based and not necessarily calibrated equivalently across compartments, so the compartment OEF^max^ contrast reflects the exported index rather than true extraction. Third, the PBZ label is compositionally mixed, containing variable admixtures of infiltrative tumour, oedema and comparatively preserved parenchyma [21, 22, 23, 24], and all volumetric features depend on segmentation quality. Fourth, DSC estimates are vulnerable to partial-volume effects at compartment interfaces and in small compartments, for which no validated glioma-specific minimum volume has been established [25, 26], and gradient-echo acquisition is sensitive to a broad range of vessel sizes [27]. Fifth, the findings describe one commercial implementation, and other MR-based approaches use different models [28]. Sixth, structural input configuration was partly confounded with acquisition era, vendor and field strength, so that comparison is observational. Last, all histologically confirmed first-diagnosis cases in the accrual window entered screening, but eligibility depended substantially on DSC and structural-sequence availability, which reflected routine protocol workflow; associated selection effects cannot be excluded.

In summary, in untreated *IDH*-wildtype glioblastoma the enhancing-versus-peritumoral perfusion phenotype was reproduced under automated whole-compartment analysis across the full six-map panel. The accompanying rCMRO₂^max^ contrast was predominantly flow-driven but dependent on lesion size. Derived oxygen-handling maps should therefore be interpreted within the complete perfusion panel and with attention to lesion size.

## Statements and Declarations

### Ethics approval

This retrospective study was approved by the institutional review board of LMU Munich (approval no. 24-0692) and was conducted in accordance with the Declaration of Helsinki.

### Consent to participate

The institutional review board waived the requirement for written informed consent for this retrospective analysis of clinically acquired imaging.

### Consent for publication

Not applicable. No individually identifiable patient information is presented; imaging in Figure 1 is shown in de-identified, defaced form.

### Author contributions

M.Ö. and J.R. contributed to conceptualisation, methodology, software, formal analysis and visualisation. M.Ö., A.N., R.S. and J.R. contributed to investigation and data curation. M.Ö., R.F. and J.R. contributed to writing of the original draft. All authors contributed to review and editing. J.R. provided supervision.

### Funding

The authors received no financial support for the research, authorship or publication of this article.

### Competing interests

R.F. has received speaker honoraria from Cercare Medical. J.R. has received travel support from Cercare Medical. All other authors declare no relationships with any company whose products or services may be related to the subject matter of this article. Cercare Medical had no data access and no role in study design, cohort selection, image processing, statistical analysis, or the decision to submit.

### Data availability

The de-identified compartment-level feature table supporting the analyses, together with the endpoint registry, is available at Zenodo (DOI 10.5281/zenodo.21908596). Individual patient imaging cannot be shared publicly under the terms of the ethics approval; requests for access should be directed to the corresponding author.

### Code availability

Analysis code is available from the corresponding author on reasonable request.

## Acknowledgements

We thank Freja Holm Etzerodt, Director of Product at Cercare Medical, for written technical clarification of the Neurosuite implementation, including the definition of OEF^max^, the rCMRO₂^max^ model relation and map-export operations. The contribution was limited to technical clarification.

## Use of AI assistance

The authors used Claude (Anthropic, Opus 5) to refine the clarity and scientific tone of the manuscript text and to enhance or check analysis code for errors. AI use was limited to language refinement and code review. Conceptualization, literature search, data interpretation, and critical reasoning were performed exclusively by the authors. All AI-assisted textual and code-related outputs were reviewed, verified, and, where necessary, edited by the authors, who take full responsibility for the accuracy, integrity, and reproducibility of the final manuscript and analyses. No AI tool was used to generate or verify references.

## Preprint

A preprint of this manuscript has been deposited on medRxiv [insert: DOI].

## Abbreviations

ADC: apparent diffusion coefficient
BCa: bias-corrected accelerated
CE: contrast-enhancing
CI: confidence interval
CTH: capillary transit time heterogeneity
DSC: dynamic susceptibility contrast
FLAIR: fluid-attenuated inversion recovery
*IDH*: isocitrate dehydrogenase
IQR: interquartile range
MTT: mean transit time
NAWM: normal-appearing white matter
OEF^max^: maximum oxygen extraction fraction
PBZ: peritumoral brain zone
rCBF: relative cerebral blood flow
rCBV: relative cerebral blood volume
rCMRO₂^max^: relative maximum cerebral metabolic rate of oxygen

## Supplementary Methods

### Acquisition

GRE-DSC perfusion used a preload-based gadobutrol protocol (0.05 mmol/kg preload; 0.1 mmol/kg bolus at 3–5 mL/s; 20-mL saline flush; preload-to-DSC delay approximately 5–6 minutes); scanner-specific sequence parameters are given in Supplementary Table S1. Post-contrast T1-weighted and T2-weighted imaging were available in all eligible cases; FLAIR and DWI/ADC were used when available.

### Eligibility

Required post-contrast T1, T2 and/or FLAIR, GRE-DSC, exported DSC maps, valid segmentation, valid NAWM reference, and analysable CE and PBZ masks. Exclusions: prior therapy, severe motion/susceptibility artefact, corrupted/incomplete DSC, segmentation failure, failed registration or map quality control, invalid NAWM, or compartment volumes below the minimum analysable volume. Missing/non-analysable necrosis did not preclude inclusion.

### Registration and NAWM normalisation

Perfusion parametric maps were rigidly registered to each patient’s post-contrast T1-weighted image using ANTs 2.6.5 with six degrees of freedom and a mutual-information similarity metric, and resampled once into the reference space using trilinear interpolation. Label maps, including the parcellations and the tumour segmentation, were resampled using nearest-neighbour interpolation. For each patient a (NAWM) mask was generated in the native T2-weighted image from the whole-brain SynthSeg 2.0 parcellation and the nnU-Net tumour segmentation in three stages. For unilateral tumours the white-matter prior comprised contralateral cerebral white matter (SynthSeg labels 2 and 41). For bilateral tumours, bilateral cerebral white matter was restricted to the tumour-free anterior third for posteriorly located tumours, the posterior third for anteriorly located tumours, and otherwise the more distant of the two terminal thirds. The prior was eroded by 2 mm and voxels within a 3-mm dilation of the ventricles were excluded. A minimum distance of 10 mm from the whole tumour segmentation, including the T2/FLAIR-abnormal tissue, was then enforced in all patients. Within the spatial prior a two-component Gaussian mixture model was fitted to z-standardised T2-weighted intensities; voxels assigned to the lower-mean component, representing normal white matter, with a posterior probability above 0.60 were retained, excluding residual T2-hyperintense oedematous or gliotic white matter. The resulting mask underwent morphological opening and hole filling, smoothing with a 0.5-mm Gaussian kernel, removal of corpus callosum voxels and of a 5-mm border around the deep grey nuclei (thalamus, caudate, putamen and pallidum) to reduce partial-volume contamination, and retention of the largest connected component only. Each parametric map was normalised within patient to its median value in the NAWM mask, and compartment-level summaries were computed as NAWM-normalised ratios.

### Quality control

Segmentation output was rated PASS/WARN/FAIL on plausibility, containment, morphology and consistency; additional modules assessed registration, NAWM-mask plausibility, map plausibility and minimum compartment volume. FAIL cases were excluded; WARN cases were retained in the primary analyses and removed in the segmentation PASS-only sensitivity analysis (n = 106). NAWM masks were visually quality-controlled using patient-specific overlays of the T2-weighted image, NAWM mask and tumour segmentation. The master seed (20260709) was fixed with the analysis plan and is recorded in the run manifest, the pipeline configuration and every analysis output.

### Feature extraction

For each compartment (CE, PBZ, analysable necrosis) and map (rCBV, OEF^max^, CTH, rCMRO₂^max^, ADC, rCBF, MTT), the median, interquartile range on both the ratio and log scales, P90, P95, Top10Mean (mean of log values at or above P90) and the geometric-mean feature (mean of log values) were extracted, with the map-specific endpoint set following the locked registry (Supplementary Table S2), in which P95 is not defined for OEF^max^ and CTH, consistent with but not proof of the voxelwise convention. Features were computed over the voxels that were finite and strictly positive within each compartment. Because the component maps derive from the same voxelwise fit, these sets coincide: the contributing voxel count was identical for rCMRO₂^max^, rCBF and OEF^max^ in every patient and compartment, so the decomposition is computed on common finite-positive voxel support. Contributing voxel counts were in fact identical across all maps in every patient and compartment, except where a map was unavailable. Upper-tail features were computed only for compartments exceeding 1 mL, approximately 100 informative voxels, below which an upper-decile summary is unstable; median and geometric-mean features were retained at the 0.5 mL case-inclusion floor. Both thresholds were derived from the compartment-volume distribution and fixed in the analysis plan before analysis. The decomposition used per-compartment geometric-mean features, the only summary that is linear over the voxel logs and on which the additive identity therefore holds; lower-tail summaries are not linear in this sense, and in this cohort the two were closely correlated (Spearman ρ = 0.90 in the enhancing compartment, 0.89 in the whole tumour). Exported maps are resampled linearly to the output grid, subject to map-specific lower floors and upper clipping, and quantised independently to 16 bits; rCBF and rCMRO₂^max^ are clipped at brain-wide 99.5th percentiles, OEF^max^ to [0, 1] and MTT and CTH to [0, 100 s].

**Extraction model and closure. In the exported and resampled maps** the relation was reproduced only up to small numerical residuals, which were retained as an explicit term in the decomposition.

### Statistics

Paired Wilcoxon signed-rank tests on log-differences; Hodges–Lehmann estimates with bias-corrected accelerated bootstrap CIs (10,000 resamples of patients, fixed seed 20260709); standardised paired effect sizes (matched-pairs rank-biserial correlation). Identity closure was evaluated both on the vendor-export grid and after registration and resampling into the common reference space, in a matched population using the same patients, segmentation masks, minimum analysable compartment volume and finite, strictly positive voxel support on both grids; because interpolation does not commute with multiplication, the analysis-grid residual exceeds the departure attributable to the export operations alone. All bootstrap intervals resample patients jointly, so that every component term, the total and their ratios are recomputed within each replicate, and component shares are ratios of the sample means of the patient-level log differences.

Scanner and vendor differences were harmonised with feature-level ComBat, following the empirical-Bayes framework of Johnson et al. [19] with the multiplicative (scale) term fixed so that only the additive location offset is estimated; the batch was defined as vendor × field strength and the fit was performed once on the locked cohort and frozen. A patient’s CE and PBZ observations entered the same fit, and no biological covariates were included. Because CE and PBZ measurements from the same patient receive the same additive batch correction, that correction cancels when CE is compared with PBZ. Empirically, the fold-changes were numerically identical before and after harmonisation (maximum absolute difference 4×10⁻¹⁶). ComBat has been applied to DSC-derived radiomic features in multicentre glioma cohorts [29] but has not been validated for physiological perfusion values, so this is a pragmatic scanner adjustment rather than an established standard for these variables.

### Sensitivity analyses

segmentation PASS-only (n = 106); no inclusion floor and a stricter 1 mL floor; unharmonised (no-ComBat) features for the cross-patient analyses; and the exploratory morphological subgroups (single-lesion cases, lesion multiplicity being confirmed by visual review of all cases flagged by the automated connected-component analysis; exclusion of lesions whose whole-tumour mask crosses the midline given their effect on the contralateral NAWM reference).

### Morphological analyses

An exploratory sensitivity estimated the three primary median-feature CE-versus-PBZ effects separately within tumour-core volume tertiles. Because the tertile results suggested variation in the magnitude of the perfusion-based effects, descriptive models regressed each primary paired log contrast on log₂ tumour-core volume and report the multiplicative change in the contrast per doubling of volume, with patient-level BCa bootstrap intervals. These analyses were outcome-blind and were not entered into a confirmatory error-rate family.

## Supplementary figures

**Supplementary Figure S1.**
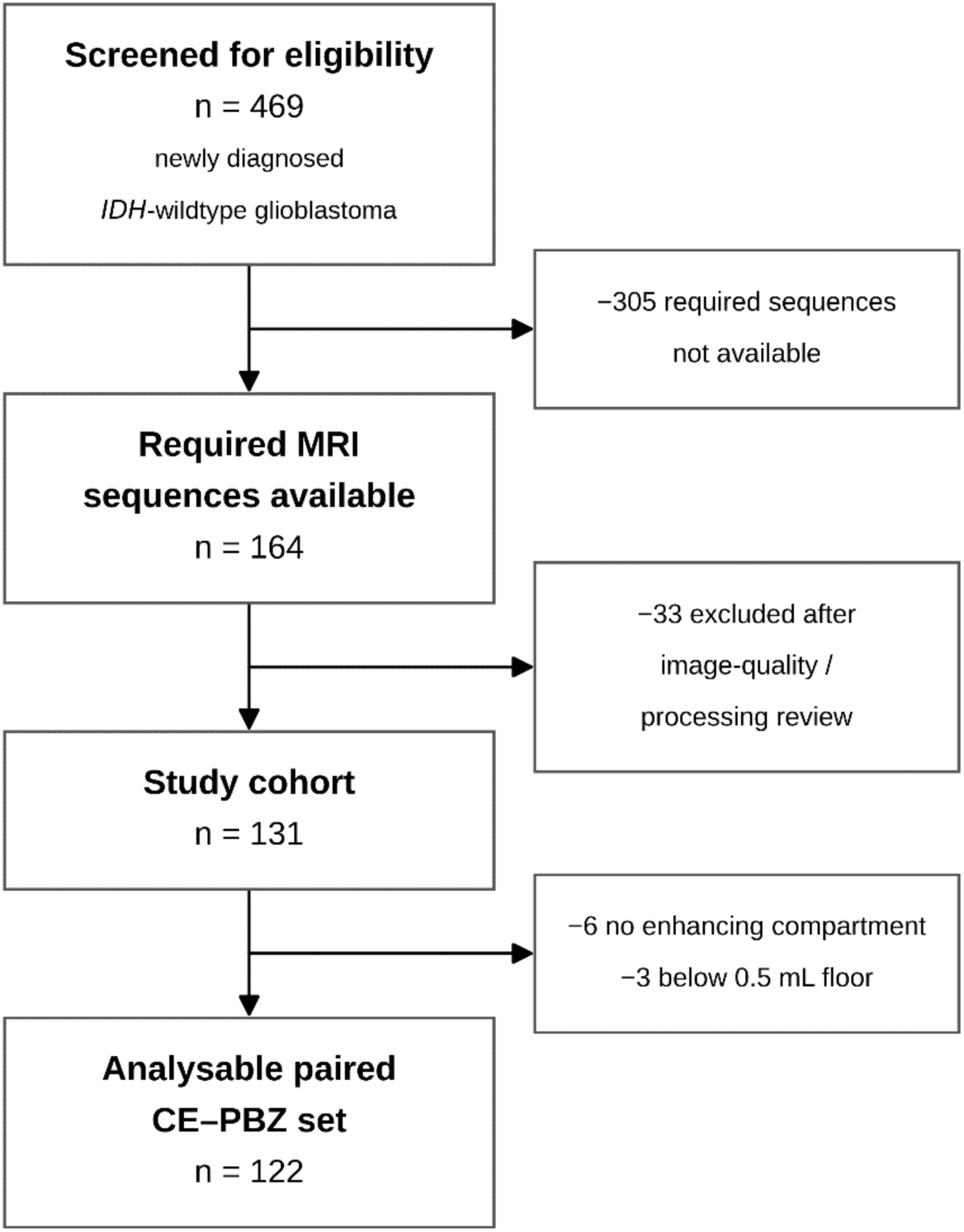
Derivation of the analysable cohort for the paired contrast-enhancing tumour (CE) versus peritumoral brain zone (PBZ) comparison. Of 469 patients screened, 305 lacked one or more of the required sequences (post-contrast T1-weighted, T2 and/or FLAIR, and gradient-echo DSC with exported parametric maps). Diffusion-weighted imaging with ADC was not required for entry and was analysable in 120 of the 122 patients in the paired set. Of the 164 remaining, 33 were excluded after image-quality and processing review: 29 for severe motion or susceptibility artefact, or corrupted or incomplete DSC series, and 4 for quality-control failure (3 segmentation, 1 registration). A further 6 patients had no enhancing compartment and 3 fell below the 0.5 mL compartment inclusion floor. Endpoint-specific samples were drawn from the final paired set (n = 122): n = 122 for the DSC-derived perfusion and oxygenation maps, n = 120 for ADC (two lacking diffusion), n = 120 for the perfusion upper-tail features and n = 118 for the ADC upper-tail features, both of which additionally required a per-compartment volume above 1 mL. DSC, dynamic susceptibility contrast.

**Supplementary Figure S2.**
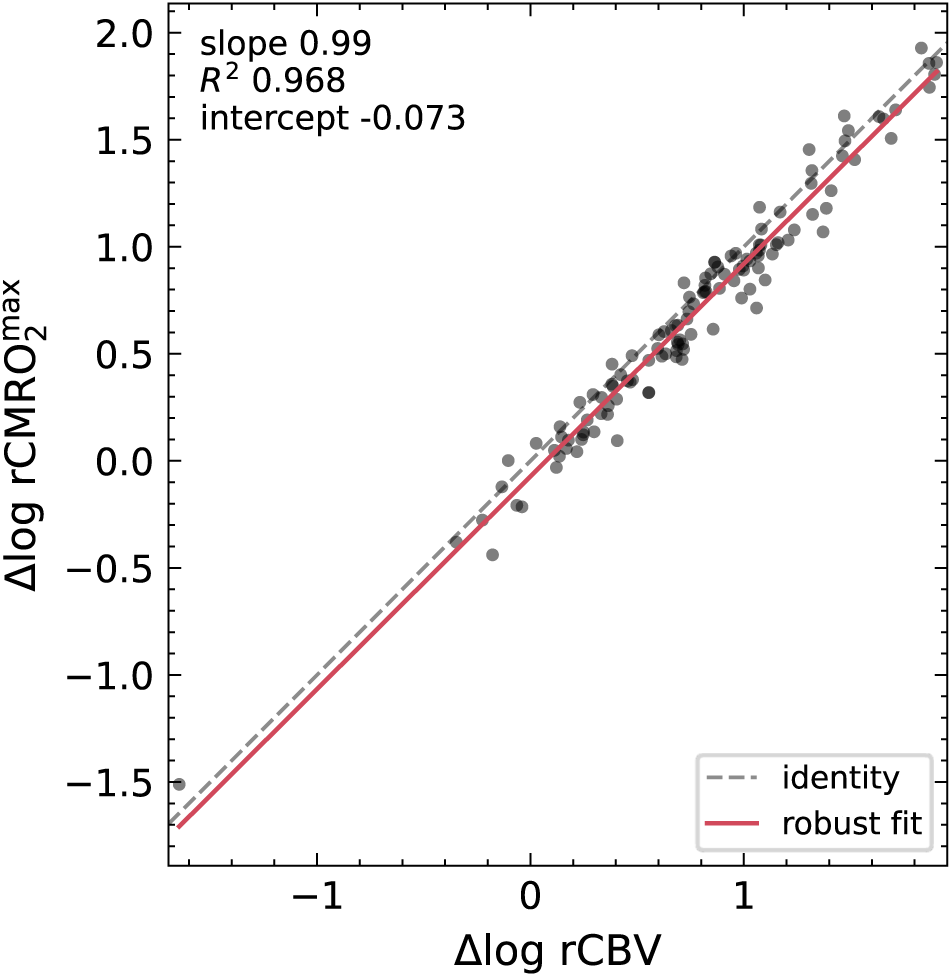
Paired Δlog rCMRO₂^max^ against Δlog rCBV (per-compartment geometric-mean features; n = 122). Each point is one patient. The grey dashed line is identity and the solid line the robust (Huber) fit (slope 0.993, intercept −0.073; ordinary-least-squares R² = 0.968). The paired compartment effect size was marginally larger for rCBV than for rCMRO₂^max^ (rank-biserial difference +0.021 [95% CI 0.007, 0.051]); this concerns compartment separation, not diagnostic performance. The intercept reconstructs exactly from the mean extraction-minus-transit terms, and the robust and ordinary-least-squares estimates were nearly identical. Because both quantities are model-linked to flow, the regression quantifies their association rather than testing physiological independence, and the slope is descriptive rather than a structural estimate: both axes are measured with error, which attenuates the fitted slope toward zero. Δlog = ln(CE/PBZ); CE, contrast-enhancing tumour; PBZ, peritumoral brain zone; rCBV, relative cerebral blood volume; rCMRO₂^max^, relative maximum cerebral metabolic rate of oxygen.

**Supplementary Figure S3.**
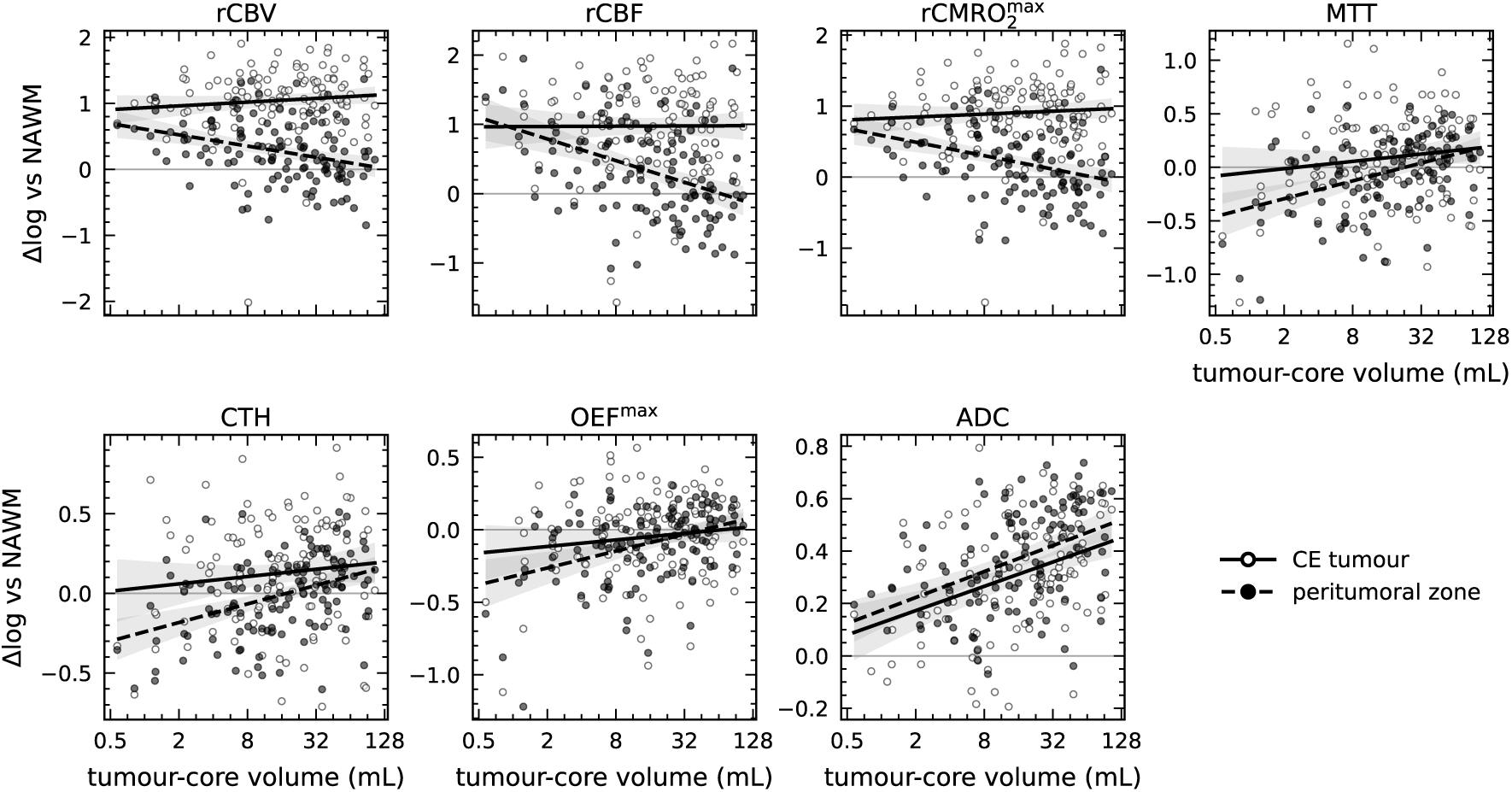
Compartment-versus-NAWM contrasts against tumour-core volume for each map. Per-compartment geometric-mean features (n = 122; ADC n = 120) plotted against tumour-core volume on a logarithmic axis. Open symbols and solid lines, contrast-enhancing tumour; filled symbols and dashed lines, peritumoral brain zone. Lines are ordinary least-squares fits with 95% mean-prediction bands. The horizontal reference at zero is equality with normal-appearing white matter. No enhancing-compartment slope differs significantly from zero for any perfusion map, whereas every peritumoral slope does; ADC changes in both compartments. Slope estimates with confidence intervals are given in Table 4.

## Supplementary tables

**Supplementary Table S1.**
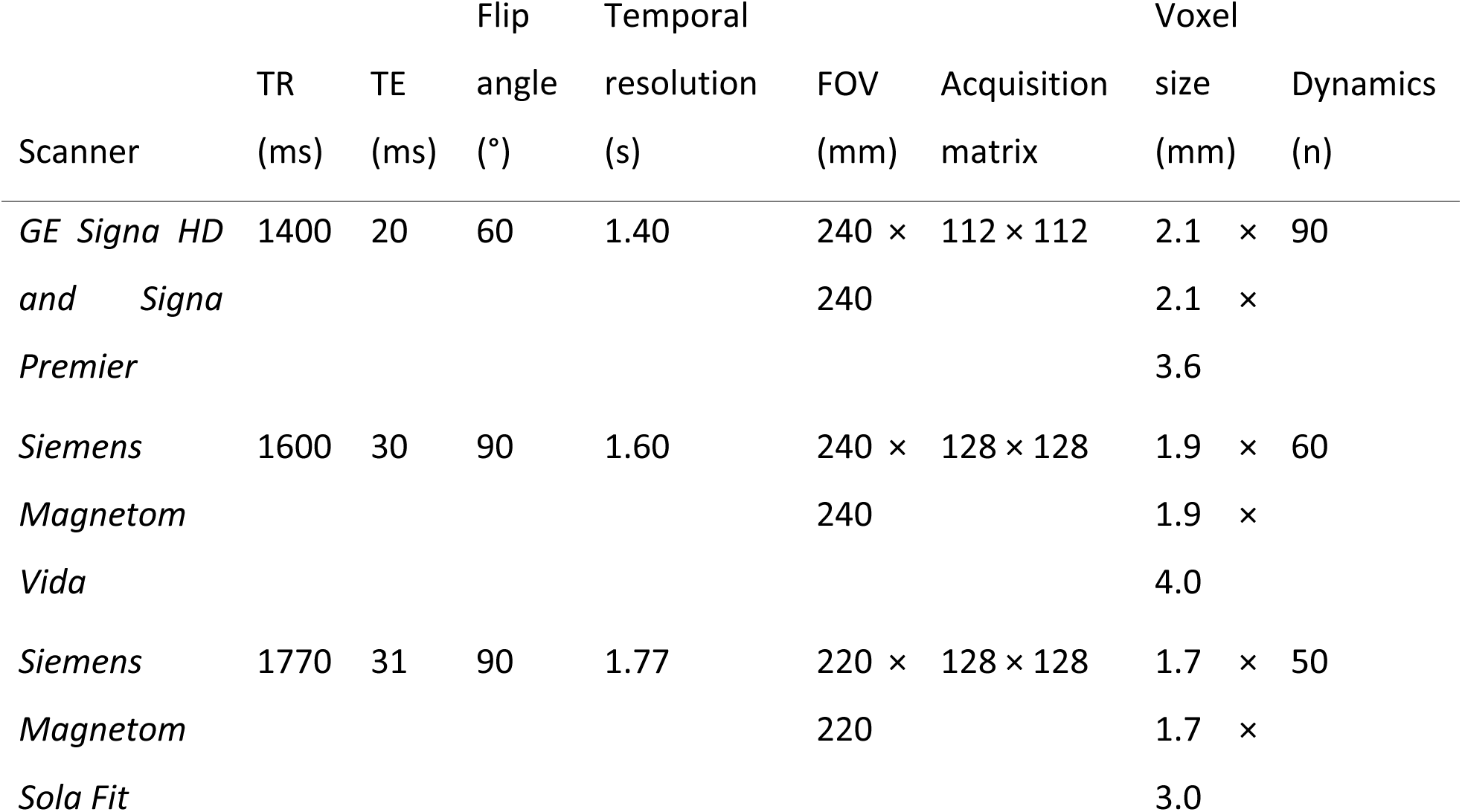
Gradient-echo dynamic susceptibility contrast (GRE-DSC) sequence parameters by scanner. All examinations used the same preload-based gadobutrol protocol described in the Supplementary Methods; the parameters below differ only in the scanner-specific sequence implementation. Voxel size is derived from the field of view and acquisition matrix together with the section thickness. Temporal resolution equals the repetition time in each implementation. Dynamics denotes the number of acquired time points, giving a first-pass acquisition of approximately 126 s, 96 s and 88 s respectively. The two GE systems used an identical DSC implementation. FOV, field of view; TR, repetition time; TE, echo time.

**Supplementary Table S2.**
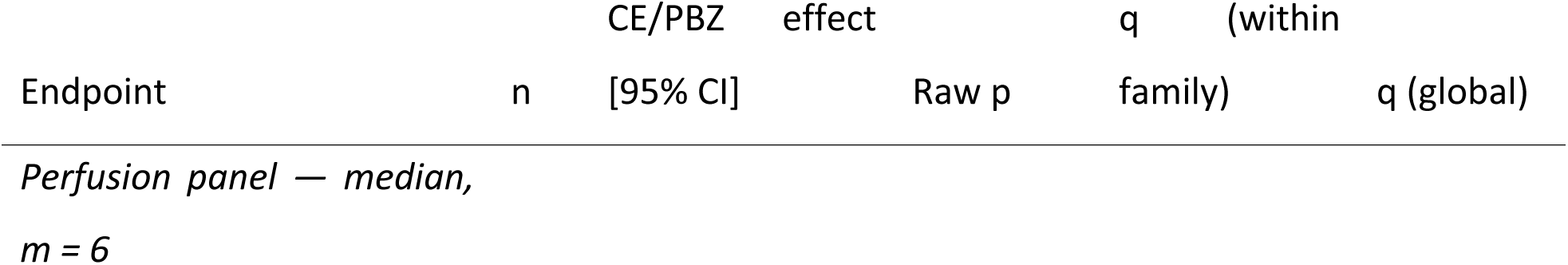
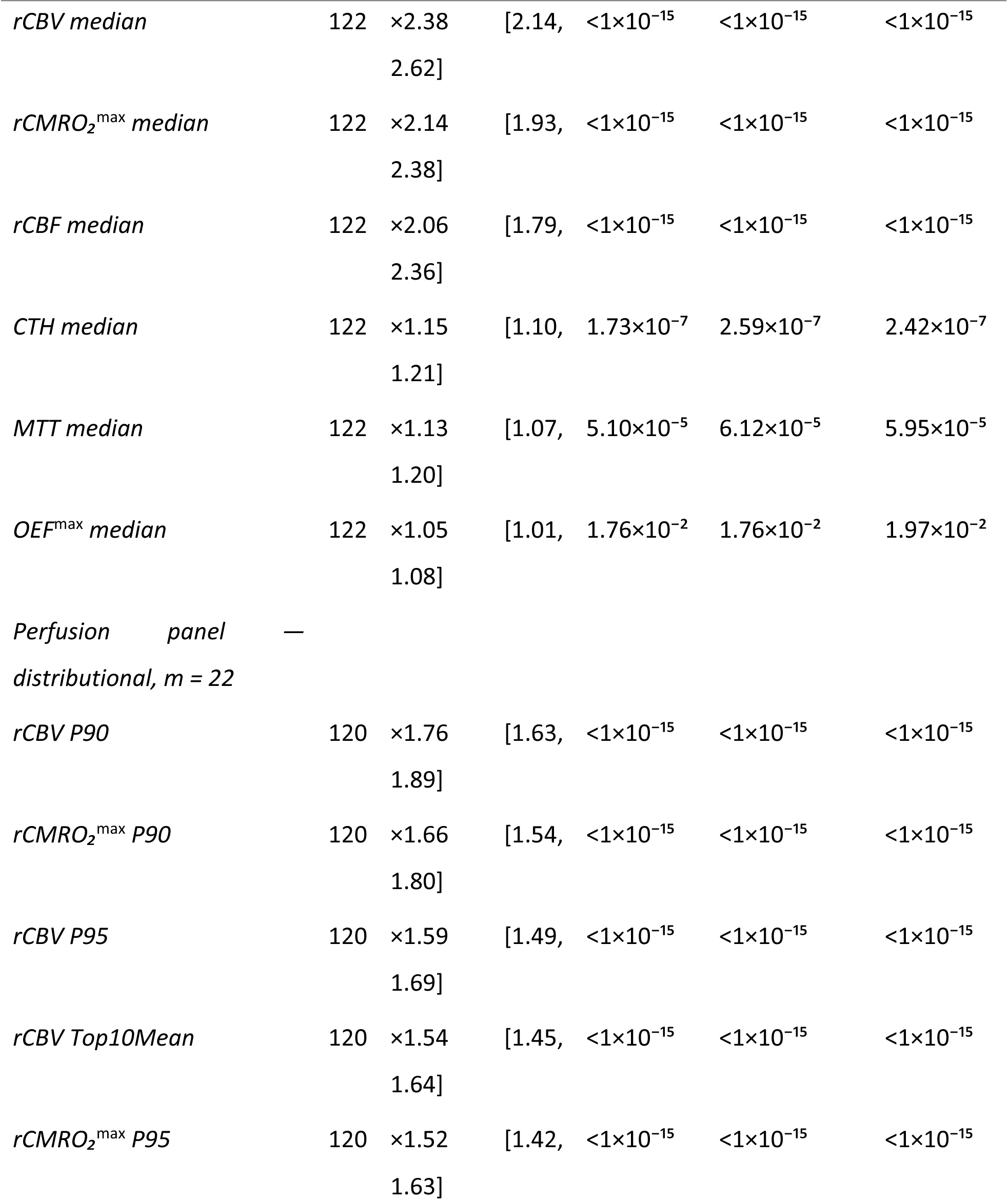
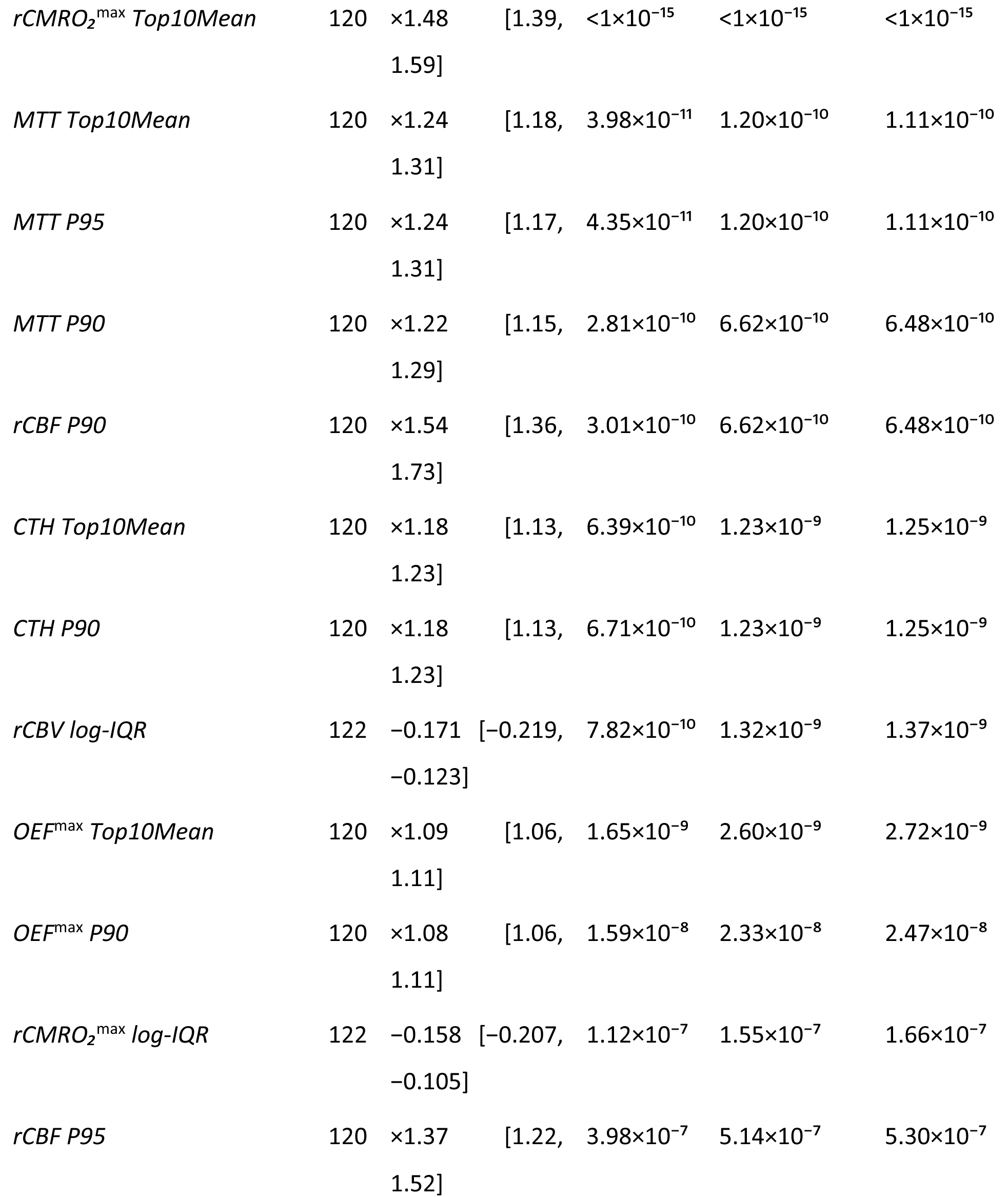
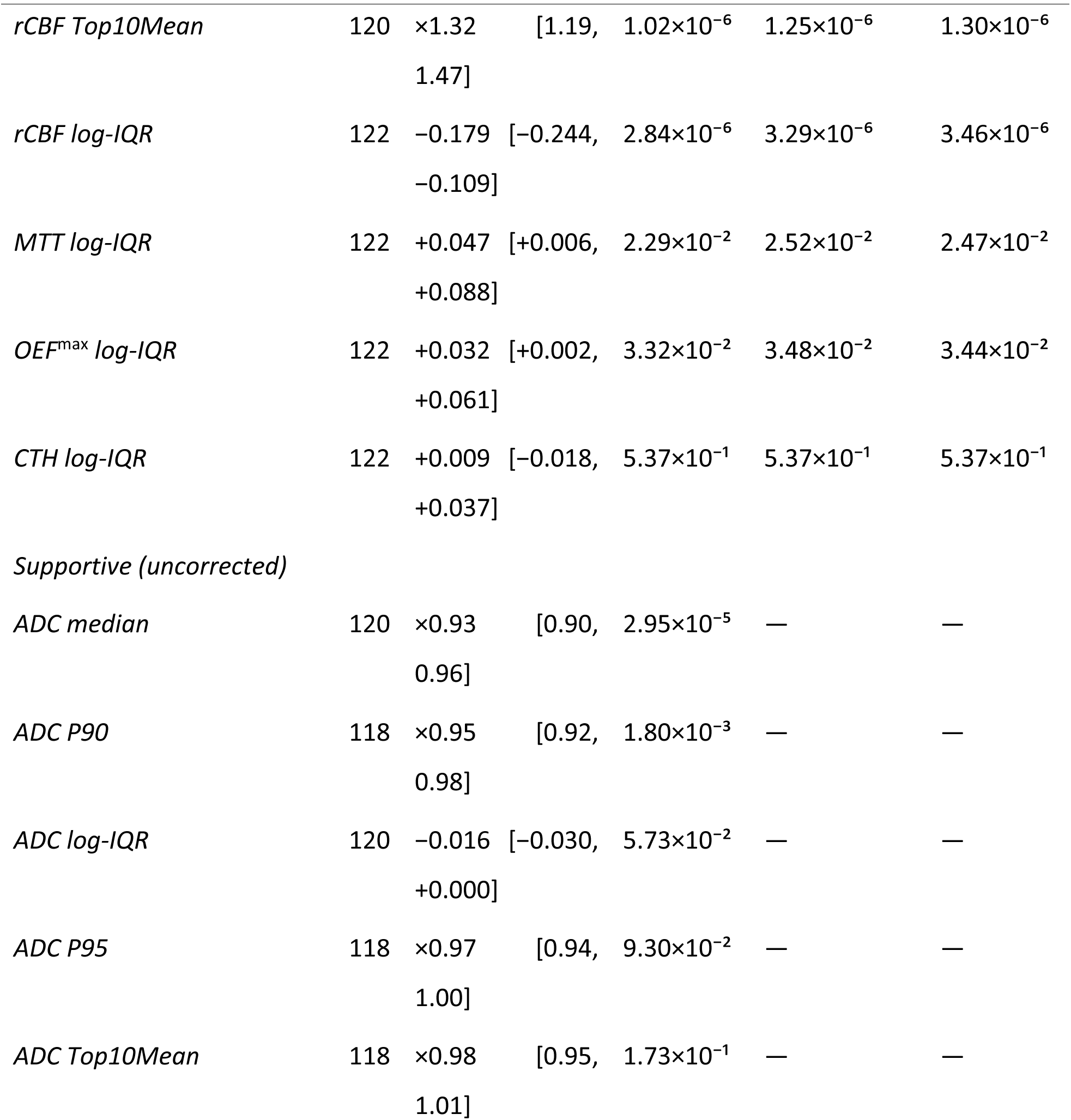
Endpoint registry, effect sizes and multiplicity control. All corrected endpoints with analysable n, the CE-versus-PBZ effect, the raw p value from the paired Wilcoxon signed-rank test, and Benjamini–Hochberg q values for the within-family correction and for the global 28-endpoint sensitivity correction. The same 27 of 28 corrected endpoints met q < 0.05 under both scopes. Location endpoints (median) are effect sizes on the median feature; distributional endpoints (P90, P95, Top10Mean, log-IQR) require an analysable compartment volume above 1 mL, giving n = 120 for perfusion and 118 for ADC. Apparent diffusion coefficient endpoints were reported as a supportive non-perfusion comparison and were not entered into either correction; the values shown for them are raw p values. Endpoint significance status was unchanged under the four-family correction structure used in the internal analysis specification, in which the three median endpoints rCBV, rCMRO₂^max^ and ADC formed one family. Effects are Hodges–Lehmann estimates with bias-corrected accelerated bootstrap 95% confidence intervals.

**Supplementary Table S3.**
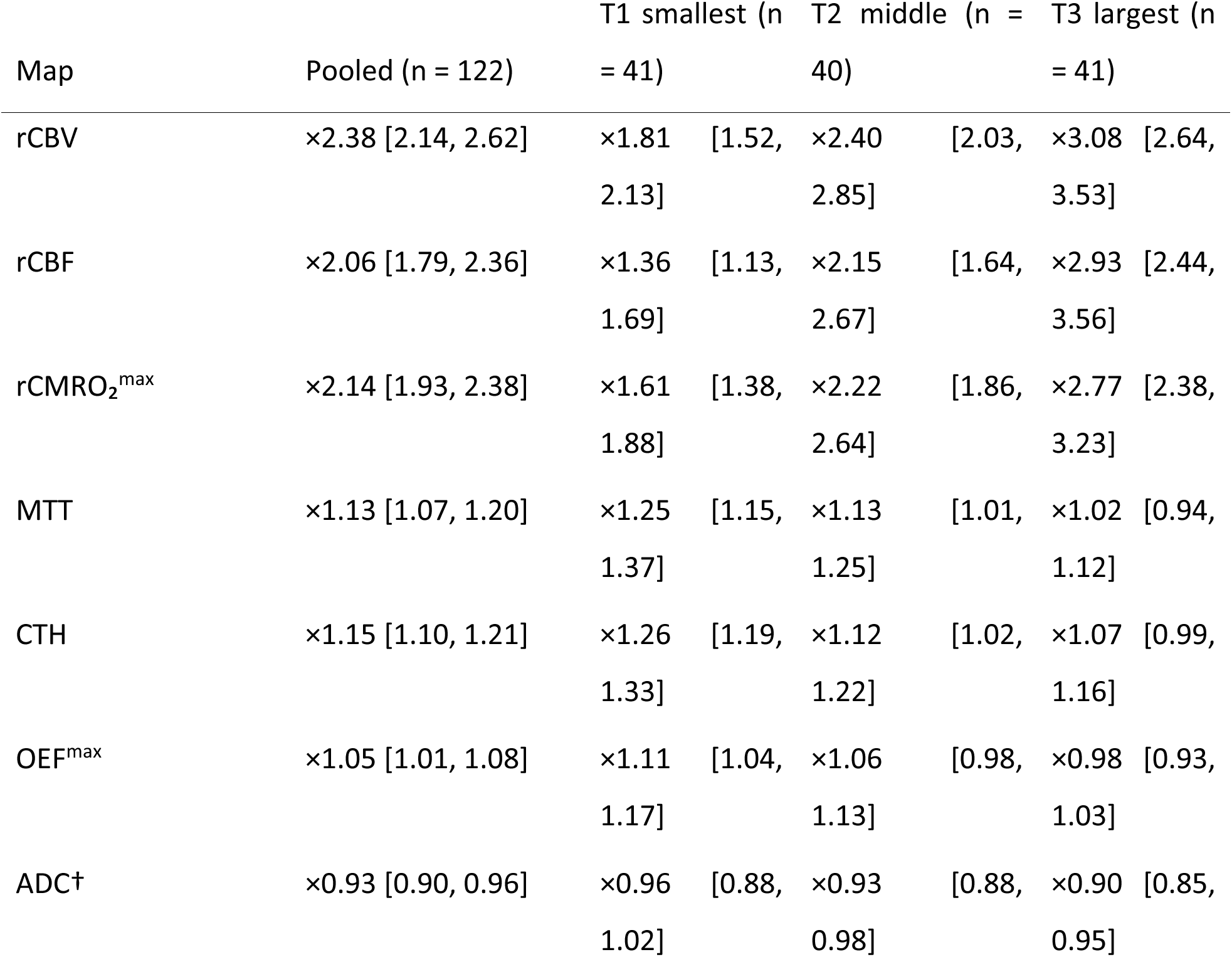
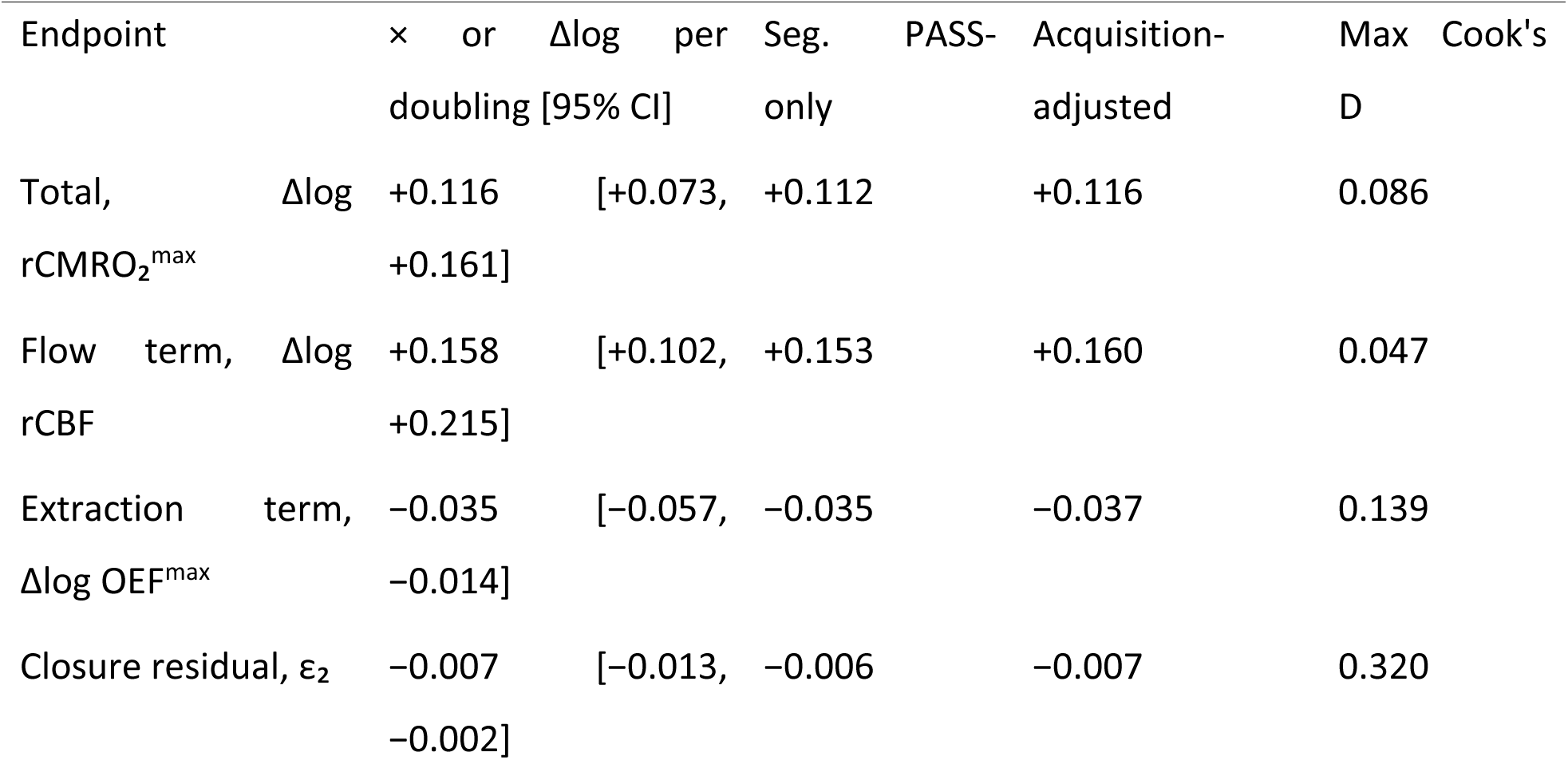
Tumour-core-volume dependence of the compartment contrasts and of the decomposition terms. Panel (a) reports CE-versus-PBZ effects within tumour-core-volume tertiles (median volume 4.3, 17.2 and 49.5 mL; n = 41, 40 and 41), as Hodges–Lehmann estimates with bias-corrected accelerated bootstrap 95% confidence intervals on median features; the pooled column repeats Table 2. Panel (b) reports the change in each decomposition term per doubling of tumour-core volume, from ordinary least-squares regression on log₂ tumour-core volume with patient-level bias-corrected accelerated bootstrap intervals (4,000 resamples, seed 20260709); the terms are given on the log scale, on which they are additive, and intervals are given for the primary estimate only. The last three columns give the same estimate in the segmentation PASS-only subset (n = 106), after adjustment for scanner vendor and field strength, and the maximum Cook’s distance as an influence diagnostic. Compartment-level slopes are given in Table 4. n = 122 throughout except ADC (120). Asterisks mark estimates whose confidence interval excludes 1. †ADC is a supportive non-perfusion comparison and was not entered into the multiplicity correction. This is an exploratory analysis. *(a) Exploratory tertile analysis* *(b) Continuous characterisation and robustness*

**Supplementary Table S4.**
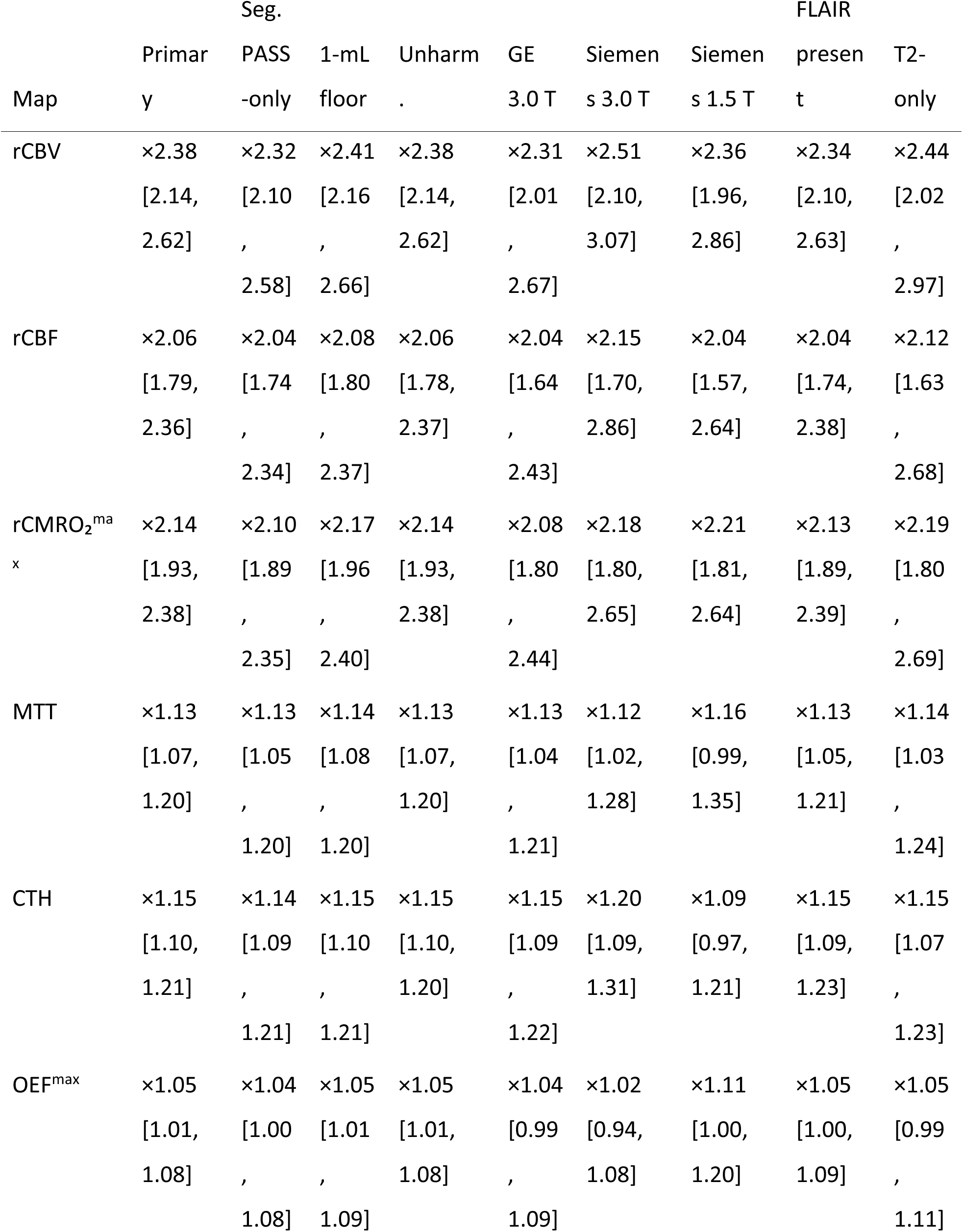
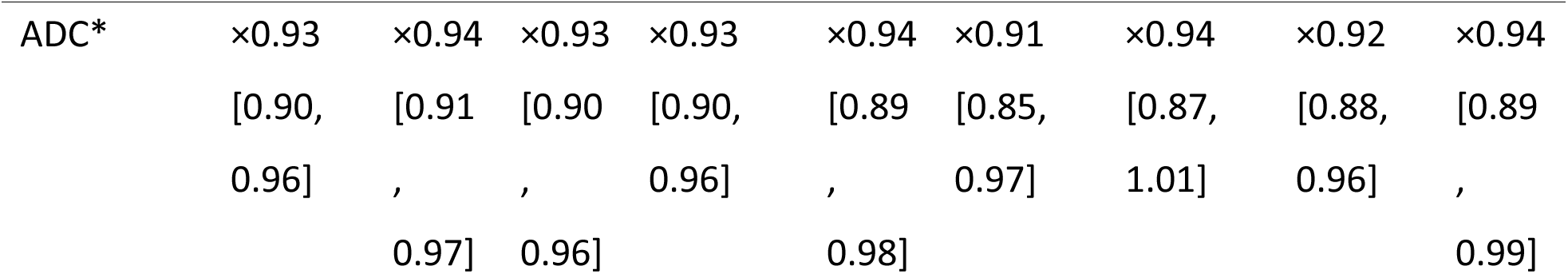
Consolidated sensitivity analyses for the six perfusion maps and supportive ADC. Paired CE-versus-PBZ effects as Hodges–Lehmann estimates with bias-corrected accelerated bootstrap 95% confidence intervals, repeated under nine conditions. Analysis choices: the primary analysis applies the 0.5-mL minimum analysable compartment volume that defines the paired set, so an unfloored analysis is not defined on this population; the 1-mL column applies the stricter floor used for upper-tail features; segmentation PASS-only (n = 106) excludes compartments with a segmentation quality-control warning; the unharmonised column repeats the analysis on the pre-ComBat extraction, and a location-only batch offset cancels in the within-patient difference, so the estimates are unchanged. Acquisition strata: GE 3.0 T (n = 65), Siemens 3.0 T (n = 33) and Siemens 1.5 T (n = 24); every stratum-specific interval overlapped the pooled estimate, and the small 1.5-T subgroup precludes inference about effect modification. Structural input: FLAIR present (n = 75) and T2-only (n = 47), with pooled flow shares of 92.8% and 92.3% respectively. n = 122 unless stated; ADC n = 120. *ADC was not entered into the perfusion multiplicity correction.

**Supplementary Table S5.**
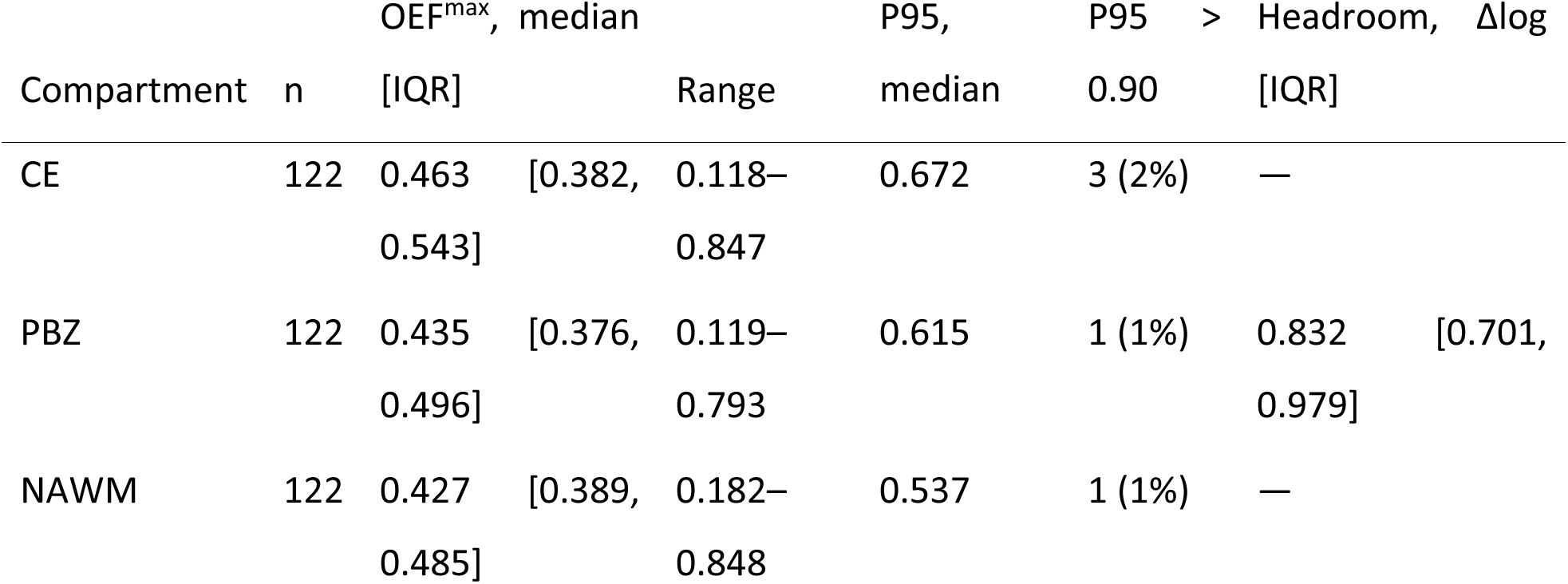
Absolute OEF^max^ by compartment and the headroom available to the extraction term (n = 122). Values are medians of the within-compartment median across patients. OEF^max^ is a fraction bounded above by one; headroom is the log distance from each patient’s PBZ value to that ceiling and is therefore defined relative to PBZ only. The observed CE-versus-PBZ extraction term (+0.046 log units, arithmetic mean of the per-patient geometric-mean feature) used 5.0% of the median headroom to the upper bound (0.904 log units, computed on the same feature). The ceiling of one is the definitional bound on a fraction, not a physiological expectation. CE, contrast-enhancing tumour; NAWM, normal-appearing white matter; OEF^max^, maximum oxygen extraction fraction; PBZ, peritumoral brain zone.

**Supplementary Table S6.**
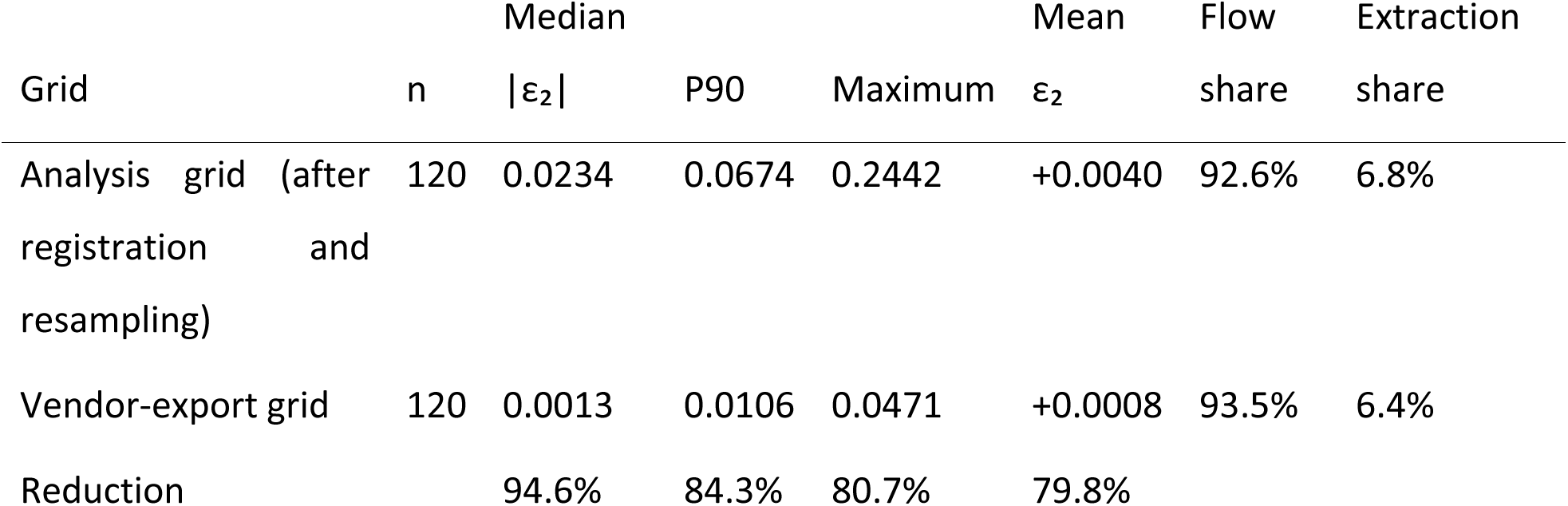
Identity closure on the vendor-export grid and after registration and resampling, in a matched population (n = 120). The residual is ε₂ = Δlog rCMRO₂^max^ − Δlog rCBF − Δlog OEF^max^ on per-compartment geometric-mean features, for which the identity is additive over voxel logarithms and the patient-specific normalisation constant cancels in the CE-versus-PBZ difference. Both rows use the same patients, segmentation masks, minimum analysable compartment volume and finite, strictly positive voxel support, and differ only in whether the maps were evaluated as exported or after registration and resampling into the structural reference space. Of the 122 paired-analysable patients, two (tumour-core volumes 0.81 and 0.57 mL) yielded no usable enhancing-compartment summary on the coarser vendor-export grid and were excluded from both rows. Component shares are computed on this matched population and differ slightly from Table 3, which reports all 122 patients on the analysis grid. Interpolation does not commute with multiplication, so the analysis-grid residual exceeds the departure attributable to the export operations alone.

## Notes

### Author Declarations

The institutional review board of Ludwig Maximilian University Munich gave ethical approval for this work.

## References

1. Boxerman JL, Quarles CC, Hu LS, Erickson BJ, Gerstner ER, Smits M, et al. Consensus recommendations for a dynamic susceptibility contrast MRI protocol for use in high-grade gliomas. Neuro Oncol. 2020;22(9):1262–1275. doi:10.1093/neuonc/noaa141.

2. Wen PY, van den Bent M, Youssef G, Cloughesy TF, Ellingson BM, Weller M, et al. RANO 2.0: update to the Response Assessment in Neuro-Oncology criteria for high- and low-grade gliomas in adults. J Clin Oncol. 2023;41(33):5187–5199. doi:10.1200/JCO.23.01059.

3. Shiroishi MS, Erickson BJ, Hu LS, Gerstner E, Hangel G, Snyder BS, et al. The QIBA Profile for Dynamic Susceptibility Contrast MRI Quantitative Imaging Biomarkers for Assessing Gliomas. Radiology. 2024;313(3):e232555. doi:10.1148/radiol.232555.

4. Jespersen SN, Østergaard L. The roles of cerebral blood flow, capillary transit time heterogeneity, and oxygen tension in brain oxygenation and metabolism. J Cereb Blood Flow Metab. 2012;32(2):264–277. doi:10.1038/jcbfm.2011.153.

5. Mouridsen K, Hansen MB, Østergaard L, Jespersen SN. Reliable estimation of capillary transit time distributions using DSC-MRI. J Cereb Blood Flow Metab. 2014;34(9):1511–1521. doi:10.1038/jcbfm.2014.111.

6. Park JE, Kim HS, Kim N, et al. Prediction of pseudoprogression in post-treatment glioblastoma using dynamic susceptibility contrast-derived oxygenation and microvascular transit time heterogeneity measures. Eur Radiol. 2024;34(5):3061–3073. doi:10.1007/s00330-023-10324-9.

7. Park YW, Han K, Jang G, Cho M, Kim SB, Kim H, et al. Tumor oxygenation imaging biomarkers using dynamic susceptibility contrast imaging for prediction of IDH mutation status in adult-type diffuse gliomas. Eur Radiol. 2025;35(11):6785–6797. doi:10.1007/s00330-025-11704-z.

8. Park YW, Choi K, Han K, Jang G, Kim H, Cho M, et al. Higher cerebral metabolic rate of oxygen derived from dynamic susceptibility contrast MRI is an independent predictor of poor survival in IDH-wildtype glioblastoma. Neuroradiology. 2026;68(11):1707–1716. doi:10.1007/s00234-026-03961-6.

9. Bonekamp D, Mouridsen K, Radbruch A, Kurz FT, Eidel O, Wick A, et al. Assessment of tumor oxygenation and its impact on treatment response in bevacizumab-treated recurrent glioblastoma. J Cereb Blood Flow Metab. 2017;37(2):485–494. doi:10.1177/0271678X16630322.

10. Reis J, Öchsner M, Adam C, Fischer TD, Liebig T, Forbrig R. Contrast enhancement is associated with a higher DSC MRI-derived cerebral metabolic rate of oxygen index in untreated glioblastoma. Diagnostics. 2026;16(9):1405. doi:10.3390/diagnostics16091405.

11. Law M, Young R, Babb J, Pollack E, Johnson G. Histogram analysis versus region of interest analysis of dynamic susceptibility contrast perfusion MR imaging data in the grading of cerebral gliomas. AJNR Am J Neuroradiol. 2007;28(4):761–766. PMID: 17416835.

12. Roques M, Raveneau M, Adam G, De Barros A, Catalaa I, Patsoura S, et al. Reproducibility of volume analysis of dynamic susceptibility contrast perfusion-weighted imaging in untreated glioblastomas. Neuroradiology. 2022;64(9):1763–1771. doi:10.1007/s00234-022-02937-6.

13. Reis J, Öchsner M, Neubauer A, et al. Peritumoral brain zone oxygen extraction fraction is associated with tumor Ki-67 index in untreated glioblastoma. Neurooncol Adv. 2026;8(1):vdag163. Published 2026 Jun 18. doi:10.1093/noajnl/vdag163.

14. Mouridsen K, Friston K, Hjort N, Gyldensted L, Østergaard L, Kiebel S. Bayesian estimation of cerebral perfusion using a physiological model of microvasculature. Neuroimage. 2006;33(2):570–579. doi:10.1016/j.neuroimage.2006.06.015.

15. Hansen MB, Tietze A, Kalpathy-Cramer J, Gerstner ER, Batchelor TT, Østergaard L, et al. Reliable estimation of microvascular flow patterns in patients with disrupted blood-brain barrier using dynamic susceptibility contrast MRI. J Magn Reson Imaging. 2017;46(2):537–549. doi:10.1002/jmri.25549.

16. Angleys H, Østergaard L, Jespersen SN. The effects of capillary transit time heterogeneity (CTH) on brain oxygenation. J Cereb Blood Flow Metab. 2015;35(5):806–817. doi:10.1038/jcbfm.2014.254.

17. Madsen LS, Thomsen MK, Angleys H, Mikkelsen IK, Brooks DJ, Eskildsen SF, et al. Estimation of oxygen extraction fraction based on haemodynamic measurements using DSC-MRI. Imaging Neurosci. 2025;3:imag_a_00562. doi:10.1162/imag_a_00562.

18. Öchsner M, Kaiser L, Stahl R, Albert NL, Liebig T, Forbrig R, Reis J. A single model for glioblastoma segmentation with and without T2-FLAIR: independent validation of a targeted dropout strategy. Front Neurol. 2026;17:1889198. doi:10.3389/fneur.2026.1889198.

19. Johnson WE, Li C, Rabinovic A. Adjusting batch effects in microarray expression data using empirical Bayes methods. Biostatistics. 2007;8(1):118–127. doi:10.1093/biostatistics/kxj037.

20. Fortin JP, Parker D, Tunç B, Watanabe T, Elliott MA, Ruparel K, et al. Harmonization of multi-site diffusion tensor imaging data. Neuroimage. 2017;161:149–170. doi:10.1016/j.neuroimage.2017.08.047.

21. Barajas RF Jr, Phillips JJ, Parvataneni R, Molinaro AM, Essock-Burns E, Bourne G, et al. Regional variation in histopathologic features of tumor specimens from treatment-naive glioblastoma correlates with anatomic and physiologic MR imaging. Neuro Oncol. 2012;14(7):942–954. doi:10.1093/neuonc/nos128.

22. Boonzaier NR, Larkin TJ, Matys T, van der Hoorn A, Yan JL, Price SJ. Multiparametric MR imaging of diffusion and perfusion in contrast-enhancing and nonenhancing components in patients with glioblastoma. Radiology. 2017;284(1):180–190. doi:10.1148/radiol.2017160150.

23. Vallatos A, Al-Mubarak HFI, Birch JL, Gallagher L, Mullin JM, Gilmour L, et al. Quantitative histopathologic assessment of perfusion MRI as a marker of glioblastoma cell infiltration in and beyond the peritumoral edema region. J Magn Reson Imaging. 2019;50(2):529–540. doi:10.1002/jmri.26580.

24. Dasgupta A, Geraghty B, Maralani PJ, Malik N, Sandhu M, Detsky J, et al. Quantitative mapping of individual voxels in the peritumoral region of IDH-wildtype glioblastoma to distinguish between tumor infiltration and edema. J Neurooncol. 2021;153(2):251–261. doi:10.1007/s11060-021-03762-2.

25. Ahlgren A, Wirestam R, Lind E, Ståhlberg F, Knutsson L. A linear mixed perfusion model for tissue partial volume correction of perfusion estimates in dynamic susceptibility contrast MRI: impact on absolute quantification, repeatability, and agreement with pseudo-continuous arterial spin labeling. Magn Reson Med. 2017;77(6):2203–2214. doi:10.1002/mrm.26305.

26. Jafari-Khouzani K, Paynabar K, Hajighasemi F, Rosen B. Effect of region of interest size on the repeatability of quantitative brain imaging biomarkers. IEEE Trans Biomed Eng. 2019;66(3):864–872. doi:10.1109/TBME.2018.2860928.

27. Stadlbauer A, Mouridsen K, Doerfler A, Hansen MB, Oberndorfer S, Zimmermann M, et al. Recurrence of glioblastoma is associated with elevated microvascular transit time heterogeneity and increased hypoxia. J Cereb Blood Flow Metab. 2018;38(3):422–432. doi:10.1177/0271678X17694905.

28. Stadlbauer A, Zimmermann M, Kitzwögerer M, Oberndorfer S, Rössler K, Dörfler A, et al. MR imaging-derived oxygen metabolism and neovascularization characterization for grading and IDH gene mutation detection of gliomas. Radiology. 2017;283(3):799–809. doi:10.1148/radiol.2016161422.

29. Manikis GC, Ioannidis GS, Siakallis L, Nikiforaki K, Iv M, Vozlic D, et al. Multicenter DSC-MRI-based radiomics predict IDH mutation in gliomas. Cancers. 2021;13(16):3965. doi:10.3390/cancers13163965.

